# Preventive oral health services and caries risk assessment of young children by non-dental healthcare providers: a scoping review

**DOI:** 10.1101/2025.09.25.25336679

**Authors:** Olubukola O. Olatosi, Robert J. Schroth, Jorma Virtanen, Carol Youssef, Nicole Askin, Moyosoreoluwa Olatosi, Julianne Sanguins, Prashen Chelikani, Josée Lavoie

## Abstract

**Background:** Early childhood caries (ECC) remains a significant global public health concern, disproportionately affecting vulnerable and underserved populations. Integrating oral health services into primary care particularly through non-dental primary care providers (NDPCPs) offers a promising strategy for early identification and prevention. However, the extent and characteristics of such integration across global health systems remain unclear.

**Objectives:** This scoping review aimed to map the available evidence on preventive oral health services (POHS) and caries risk assessment (CRA) delivered to children under six years of age by NDPCPs. Specifically, it addressed (1) the types and characteristics of POHS and CRA provided, and (2) how commonly these services are implemented in pediatric primary care settings.

**Methods:** The review followed the Joanna Briggs Institute (JBI) Scoping Review Framework. A comprehensive search of four databases, MEDLINE, EMBASE, CINAHL, and Web of Science was conducted for English-language studies published between 2009 and 2024. Eligible studies included primary research on CRA, fluoride varnish application, dental referrals, or oral health promotion delivered by NDPCPs (e.g., physicians, nurses, dietitians) to children under six. A total of 54 studies met the inclusion criteria and were charted, summarized, and analyzed narratively.

**Results:** Most studies (83%) were conducted in the United States, with additional representation from Australia, the United Kingdom, Europe, and limited low– and middle-income countries (LMIC) settings. Interventions were delivered by a wide range of NDPCPs including physicians, nurse practitioners, and health visitors, often during well-child visits. Common interventions included oral health education, CRA using structured tools, fluoride varnish application, and dental referrals. Several studies reported improved service uptake following provider training, electronic medical record (EMR) integration, or Medicaid reimbursement policies. Despite evidence of effectiveness and feasibility, no eligible studies were identified from e.g. Canada, highlighting a critical implementation gap.

**Conclusions:** NDPCPs play an important and increasingly well-supported role in the delivery of POHS to young children. This review underscores the need for policy frameworks such as reimbursement mechanisms and interprofessional training to support oral health integration into primary care. Canada may benefit from adopting system-level policies to enable and evaluate CRA and POHS in primary care, especially for underserved and Indigenous populations.

## Introduction

Early childhood caries (ECC) is a highly prevalent, yet preventable, disease that affects millions of children globally, with particularly high rates among socioeconomically disadvantaged and underserved populations. Defined as the presence of one or more decayed, missing, or filled tooth surfaces in any primary tooth of a child under six years of age, ECC has significant health, developmental, and psychosocial consequences (Colak et al., 2013; Tungare & Paranjpe, 2025). These include pain, infection, impaired nutrition and growth, difficulties with speech and learning, and reduced quality of life. ECC also imposes considerable burdens on health systems, often requiring costly treatment under general anesthesia (Schroth et al., 2016).

Timely prevention and early detection are critical to minimizing the impact of ECC. Recognizing this, several professional organizations including the United States Preventive Services Task Force (USPSTF), the American Academy of Pediatrics (AAP), and the Canadian Pediatric Society (CPS) recommend the integration of oral health promotion and caries prevention into routine pediatric care (Davidson et al., 2021; Holve et al., 2021; Krol & Whelan, 2023). These recommendations include the application of fluoride varnish, oral health screening, CRA, anticipatory guidance, and dental referrals during well-child visits. NDPCPs including pediatricians, nurses, family physicians, and nurse practitioners frequently interact with children in early life and are well-positioned to deliver these POHS (Gaffar et al., 2023; Quinonez et al., 2014).

Despite these policy-level endorsements, the uptake and implementation of CRA and related services by NDPCPs remains limited in many settings (Harnagea et al., 2017; Kalhan et al., 2020; Olatosi et al., 2025b). Barriers include lack of training, limited time during office visits, uncertainty about scope of practice, and inadequate referral systems to dental care providers (Gaffar et al., 2023; Olatosi et al., 2025a; Rabiei et al., 2014). These gaps contribute to continued disparities in oral health outcomes, particularly in communities with limited access to dental professionals. Embedding CRA and oral health services within primary care aligns with calls for integrated, equity-oriented approaches to child health and reflects broader shifts toward interprofessional collaboration in healthcare delivery (Lienhart et al., 2023; Olatosi et al., 2025c).

This scoping review was guided by a health equity lens, recognizing that ECC disproportionately affects children from low-income, Indigenous, and rural communities who face structural barriers to accessing timely dental care (Kyoon-Achan et al., 2021; Rowan-Legg & Canadian Paediat, 2013). Integrating CRA into primary care settings represents a pragmatic strategy to reduce these inequities by leveraging existing health system touchpoints. Moreover, initiatives such as the “Smiles for Life” curriculum have sought to train NDPCPs in the delivery of POHS (Douglass et al., 2007).

## Objectives

The purpose of this scoping review was to map the available evidence on POHS and CRA provided to children under six years of age by NDPCPs. The review seeks to address the following research questions:

1. What is the evidence of POHS and CRA for young children by NDPCPs?
2. How common is the practice of POHS and CRA for this population?

This review aims to identify the scope, characteristics, and outcomes of such interventions, and highlight gaps in knowledge to inform future research, policy, and implementation strategies for oral health integration into primary care.

## Methods

This scoping review was conducted in accordance with the methodology recommended by the Joanna Briggs Institute (JBI) Scoping Review Framework. It draws on the original framework developed by Arksey and O’Malley (Arksey & and O’Malley, 2005), later enhanced by Levac et al (Levac et al., 2010). The JBI framework outlines six key stages: (1) identifying the research questions; (2) identifying relevant studies; (3) selecting studies; (4) charting the data; (5) collating, summarizing, and reporting the results; and (6) consulting stakeholders.

### Stage 1: Identifying the Research Questions

This review was guided by the following research questions:

1. What is the evidence of CRA and POHS for young children by NDPCPs?
2. How common is the practice of CRA and POHS for this population?

### Stage 2: Identifying relevant studies

Relevant studies were identified through a comprehensive search of four electronic databases: MEDLINE (Ovid), EMBASE (Ovid), CINAHL with Full Text (EBSCO), and Web of Science Core Collection (Clarivate). The search was conducted May 2024. A modified version of the Children Filter (Broad) from MEDLINE was used in combination with subject headings and keyword terms representing concepts of preventive dental care and non-dental health professionals. The search was limited to English-language publications from 2009 onward. The initial MEDLINE strategy was developed in collaboration with a health sciences librarian at the University of Manitoba and peer-reviewed using the Peer Review of Electronic Search Strategies (PRESS) checklist (Canada’s Drug Agency, 2016). The final search strategies for all databases are presented in **Appendix 1**.

### Stage 3: Study Selection

Study selection was performed in two phases: title and abstract screening, followed by full-text review. Studies were included if they met the following criteria:

#### Inclusion criteria

- Published in English from 2009 onward.
- Primary research studies (e.g., observational or interventional designs) excluding reviews.
- Focused on CRA, dental screening, oral health promotion, fluoride varnish application, dental referral, or other POHS delivered by NDPCPs to children under six years of age.
- Involved NDPCPs, including nurses (e.g., registered nurses, licensed practical nurses, and nurse practitioners), physicians (e.g., family physicians, pediatricians), pharmacists, physician assistants, dietitians, nutritionists, physical therapists, speech-language pathologists, and trainees in these professions.

#### Exclusion criteria

- Studies focused solely on children older than six years.
- Studies involving only dental professionals.
- Studies involving lay health workers (community members without formal health professional training).
- Studies assessing only knowledge, attitudes, or self-reported practices without evaluating actual service delivery.
- Review articles, editorials, commentaries, opinion pieces, and articles without full-text availability.

A total of 9,757 references were retrieved and imported into EndNote for deduplication. After removing 4,538 duplicates, 5,219 records were screened using Covidence. Title and abstract screening were conducted by the primary reviewer and six trained researchers. Articles with uncertain eligibility were advanced to full-text review. Four reviewers, including the primary reviewer, conducted the full-text screening. Discrepancies were resolved through consensus discussions. A total of 54 studies met the inclusion criteria. The study selection process is summarized in the Preferred Reported items in Systematic Reviews and Meta-analysis (PRISMA) flow diagram(Moher et al., 2009) (Figure 1).

**Figure 1:**
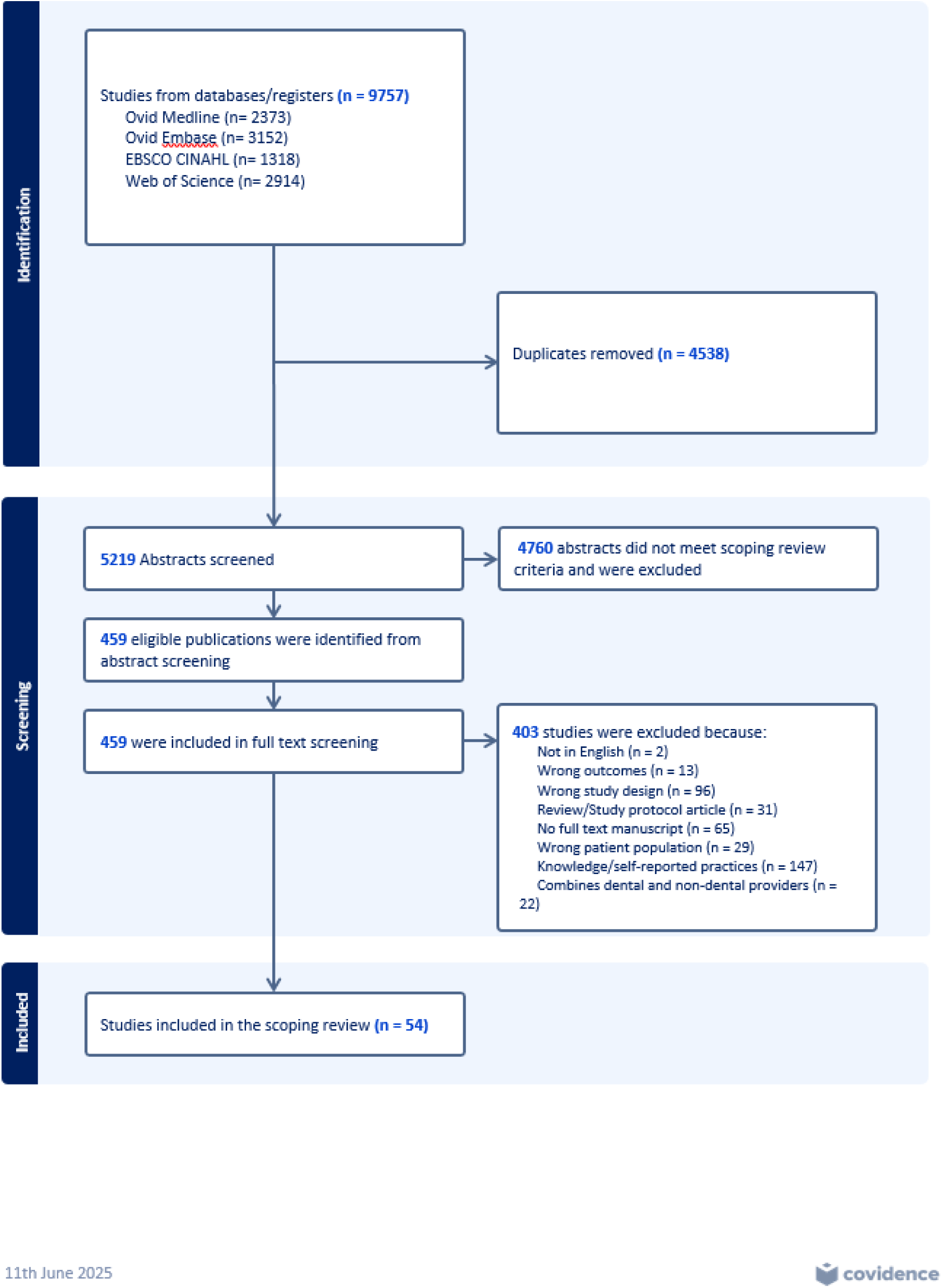
PRISMA Flow Chart for the Scoping Review.

### Stage 4: Charting the Data

Data extraction was conducted using Covidence, guided by a standardized extraction template. The following data were extracted from each included study:

- Author(s) and year of publication
- Country of study
- Study aim
- Setting
- Study design
- Participant characteristics (e.g., age)
- Type of healthcare provider
- Inclusion and exclusion criteria
- Description of the intervention(s)
- Reported outcomes
- Key findings

### Stage 5: Collating, Summarizing, and Reporting the Results

A narrative synthesis of the findings was undertaken. Results were summarized descriptively, highlighting the scope, characteristics, and outcomes of CRA and preventive oral health interventions delivered by NDPCPs. Key themes were identified through content analysis, and research gaps were noted to inform future directions for integrating oral health into pediatric primary care.

## Descriptive analysis

### Overview of included studies

This scoping review identified 54 studies published between 2009 and 2024 that met the inclusion criteria. Most of the studies (n = 45, 83%) were conducted in the United States, with the remaining studies originating from Australia (n = 2) (Maher et al., 2012; Neumann et al., 2011), the Netherlands (n=1) (Verlinden et al., 2024), Sweden (n=1) (Brännemo et al., 2021), Scotland (n=1) (Turner et al., 2010), Cambodia (n=1) (Turton et al., 2021), the Dominican Republic (n=1) (Abreu-Placeres et al., 2023), Norway (n=1) (Wigen & Wang, 2017), and Peru (n=1) (Melgar et al., 2024) (Figure 2). Study designs varied and included retrospective cohort analyses, cross-sectional surveys, quasi-experimental and randomized controlled trials, quality improvement initiatives, and observational studies. Sample sizes ranged from fewer than 50 participants to more than 2 million children in large administrative datasets (see Table 1).

**Figure 2:**
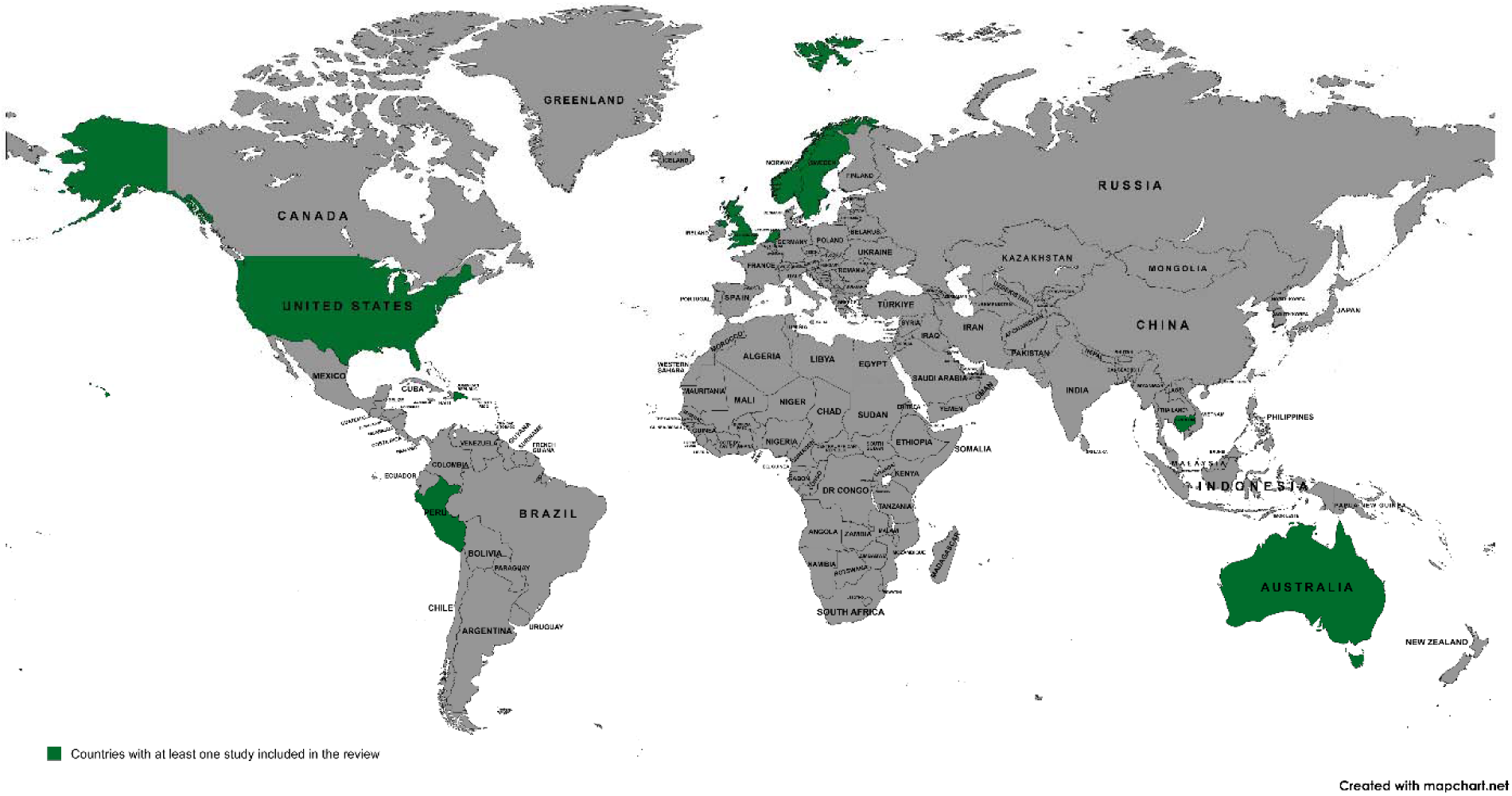
Countries with at least one study included in the review.

**Table 1.** Characteristics of included studies examining CRA and POHS delivered by NDPCPs to children under six years of age (n=54). This table summarizes study design, provider type, interventions, and key findings.

| Author (Year):<br>Country | Study Aim | Study Design | Population<br>Age/Sample<br>size | Provider<br>Type | Intervention(s) | Main findings |
| --- | --- | --- | --- | --- | --- | --- |
| Meyer and Danesh<br>(2021): USA | To compare FV utilization rates at dental visits and WCV using data from the Medicaid programs in Ohio and North Carolina | Retrospective | 1–5 yrs. | Physician | FV | During the pandemic, the quarterly fluoride utilization rate significantly ↓ both at medical WCV and dental visits. |
| McCulley et al.<br>(2022):<br>USA | To further develop and psychometrically test a de novo instrument on academic social bullying with health sciences educators for content and construct validation | Retrospective<br>+ Ed. | 6 mo – 5<br>yrs.<br>129 children | Physician,<br>NP, MA | Education,<br>Screening,<br>CRA, FV,<br>referrals | There was 94.6% improvement in oral health risk assessments, 14.7% ↑ in children identified as high caries-risk, 30.2% ↑ in oral health anticipatory guidance, and 100% ↑ in dental referrals. |
| Kranz et al.<br>(2022):USA | The study examined the association between Medicaid payment and receipt of FV during pediatric medical visits | Retrospective | 6 mo – 5 yrs<br>3,393,638<br>medical<br>visits | Pediatric<br>Medical<br>Visits | FV | Fewer than one in 10 visits included FV. ↑Medicaid payment was positively associated with receipt of FV during pediatric medical visits. Attention should be paid to the effects of provider payment on access to pediatric oral health services. |
| Okunseri et al.<br>(2009): USA | To determine the extent by which a state-level policy change impacted access to FVT for Medicaid enrolled children | Claims<br>Analysis | 1–6 yrs<br>3,631<br>children | Medical Care<br>Provider | FV | Following the policy change, claims for FVT ↑. FVT rates ↑ for children of both sexes and all ages. Overall, 48.6% of the ↑ in FVT was attributable to medical care providers. The largest ↑ was seen in children 1 to 2 yrs, among whom medical care providers were responsible for 83.5% of the ↑. |
| Pahel et al.<br>(2011):USA | To estimate the effectiveness of a medical office-based preventive dental program IMB, which included FV application, in reducing treatments related to dental caries. | Population-based cohort study | 6 mo – 6 yrs | Physician | Education, Screening, CRA, FV, Referral | The IMB program ↓ caries-related treatments for children with 4 IMB visits. Multiple applications of FV at the time of primary tooth emergence was most beneficial. Referrals to dentists for treatment of existing during IMB implementation limited the cumulative ↓ in caries-related treatments but also contributed to improved oral health. |
| Rozier et al.<br>(2010):USA | To examine the (1) delivery of POHS in NC medical offices, in total and in relation to dentist visits; (2) factors associated with use of medical oral health and dentist visits; and (3) the effect of the medical office-based program on use of dentist services for children up to 3years of age during the first seven years of the program | Retrospective | 6 mo – 3 yrs<br>11 million child-month records for 629,005 Medicaid-enrolled children | Physician | POHS | Medicaid claims from 2000 to 2006 shows the program substantially ↑ POHS. By 2006 approximately 30% of WCV for 6- to 36-month-old children included these services. Additional strategies are needed to ensure preventive oral health care for more low-income children. |
| Berger et al.<br>(2014):USA | The article describes an oral health preventive pilot program, including the application of FV, in a Missouri rural health clinic and presents findings related to oral health screenings, epidemiology, and risk factors. | Cross-sectional | 0–5 yrs<br>36 children | NP | Education, Screening, CRA, FV, referral | NPs implemented Oral screening and FV on children 0-5 years in rural health clinic. 75% were in the high-risk category. Most of the children 97.2% had never been to the dentist. 27.8% had ECC, 25% needed urgent referral for extensive caries. |
| Kranz et al.<br>(2014b):USA | The study examined the association between who (PCP, dentist, or both) provides these services to Medicaid enrollees before age 3 years and oral health at age 5 years. | Retrospective | <3 yrs<br>5,235 children | PCP, Dentist | POHS | Most children (75%) received POHS during only IMB visits, 13% received the recommended 4 or more IMB visits before 3<sup>rd</sup> birthday. Children with multiple PCP or dentist visits had a similar number of mean dmf primary teeth in kindergarten, children with only PCP visits had a ↑ proportion of untreated decayed teeth. |
| Sudhanthar et al.<br>(2019):USA | To increase oral FV application for children starting at 6 months or the time of tooth eruption up to 3 years of age by at least 50% over 18 months | Cohort | 6 mo – 3 yrs<br>50 children | Physician, Nurse, Medical Assistant | Education, CRA, FV | FV rate ↑ from 14% to 55%, 18 months after the intervention was implemented. There was 5x ↑ in FV rate in PDSA cycle 3. FV did not affect the flow of the patients in busy primary care practice. The rate of improvement was across all the age groups, providers and in both clinical sites. |
| Cheng et al.<br>(2019):USA | To describe changes in oral health behaviors following implementation of a nursing intervention targeting children at risk for ECC at an urban 2-site primary care practice. | Retrospective | 9 mo – 5 yrs<br>2,097<br>children | Nurse | Education,<br>CRA,<br>Examination,<br>Referrals, FV | Brushing teeth $\geq 2x$ daily ↑ from 42.5% to 60.8%, using fluoride toothpaste ↑ from 50.7% to 83.4%. Improvements were noted across the 3 oral health behaviors (brushing 2x daily, use of FL toothpaste, and adult help with brushing among children <18 months). Nursing interventions show promise for promoting POHS in primary care settings. |
| Patton and Severe<br>(2020):USA | To evaluate the relationship between children's tooth decay risk score and a dental examination and parent reports of oral health practices | Cross-sectional | 456<br>Preschoolers | Nursing<br>Student | CRA,<br>Screening | The children received an oral health assessment, 21% were referred for further evaluation and dental treatment. There was low tooth brushing adherence as reported by parents. |
| Okunseri et al.<br>(2010):USA | To examine the association of rates of FVT for children enrolled in Wisconsin Medicaid with race/ethnicity, urban influence codes (UIC), and dental health professional shortage area (DHPSA) designation based on county of residence. | Retrospective | 1–6 yrs | Physician | FV | ↑ Access and utilization of FVT, substantial racial/ ethnic and geographic variation in the provision of FVT for children enrolled in Medicaid. Post-policy, the largest ↑ were observed for Native Americans residing in non-DHPSA counties, enrollees living in rural counties, and for Hispanics living in partial and entire DHPSA counties. African Americans residing in partial DHPSA and metropolitan counties displayed the lowest rates of FVT claims. |
| Pahel et al.<br>(2010):USA | To assess (1) the feasibility of matching Physician-completed encounter forms (EFs) to Medicaid claims (2) the agreement on the frequency of preventive dental visits in these two data sources and identify child and medical practice characteristics that are associated with agreement in the two data sources. | Retrospective | <3 yrs<br>41,252 EFs<br>for 30,606<br>children and<br>40,909<br>claims for<br>27,607<br>children<br>being<br>available to<br>be matched. | Physician | Education,<br>screening,<br>CRA, FV,<br>referral | Increasing age of child and residence in same county as the medical practice increased the likelihood of a match. Pediatric practices provided most visits (82.4%) and matches. |
| Taylor et al.<br>(2014):USA | Describes the Kellogg Ohio WIC Oral Health Model which provides one solution to the critical need for pediatric oral health preventative services by utilizing allied health professionals to bridge the gap | Cross-sectional | 0–5 yrs<br>4,091 | Dietitian | Education,<br>Screening,<br>CRA, FV,<br>referral | 4091 children received oral health services, mean age of children served was 2.3years, 6,655 oral health visits, and 90% of caregivers reported satisfaction with health care services. |
| Biordi et al.<br>(2015):USA | The study provided oral health care services at 2 sites using a nurse practitioner–dietitian team to increase dental workforce capacity and improve access to care for low-income preschool children. | Cross-sectional | <5 yrs<br>4,360<br>children | NP, Dietitian | Education,<br>Screening, FV | Children received FV in 7195 total visits. The number of caries ↓ with ↑ program visits, which coincided with an increase in the proportion of participants visiting a dentist. |
| Jackson<br>(2015):USA | To (a) increase the identification of primary care pediatric patients at high-risk for development of ECC and (b) effectively refer those patients to a dental provider. This process improvement project involved implementation of a risk assessment tool at the 9-, 12-, and 18-month well child visit. | QI Project | 9–18<br>months<br>106 children | Physician,<br>NP | CRA, Referral | ↑ in the identification of patients at high risk for ECC. Practice improvement intervention such as this may succeed better in sites where practitioners are motivated to implement universal screening tool to improve management of patients at-risk for ECC. |
| Murphy and<br>Larsson<br>(2017):USA | To integrate and evaluate a pediatric oral health project in an AI, pediatric primary care setting | Descriptive | 0–5 yrs<br>47 children | NP | Education,<br>CRA, Referral | In less than 5 min per appointment, the primary care provider integrated oral health screening, education, and referral into WCVs. Oral health is part of total health and thus should be incorporated into routine WCVs. |
| Braun et al.<br>(2017):USA | To assess an oral health promotion (OHP) intervention for medical providers' impact on ECC | Quasi-experimental | <3 yrs<br>4,855<br>children | Physician,<br>NP,<br>Physician<br>Assistant | Screening,<br>CRA, FV,<br>referral | OHP intervention targeting medical providers ↓ ECC when children received ≥ 4 FVAs at a medical visit by age 3 y. There was ↑ children < 3 years who received FVA at WCV. |
| Maher et al.<br>(2012):Australia | The study evaluated the degree to which child health professionals were incorporating oral health promotion and prevention into their routine care through the New South-Wales (NSW) ECOH program. | Mixed<br>methods | <5 yrs<br>3440<br>children | Child and<br>Family<br>Health Nurse | Education,<br>screening,<br>Referral | Parents received oral health information, education and support through written resources and contact with child health professionals. Child and family health nurses reported routinely incorporating oral health promotion and early identification for ECC into their practices. ↑ Referral 5x over 2 years. |
| Goldstein et al.<br>(2022):USA | Study examined whether state-level policies requiring pediatric medical providers to obtain oral health training prior to delivering POHS in medical offices increased receipt of POHS in medical offices and overall receipt of POHS in both medical and dental offices | Observational study | Children <6 years<br>2,973,171 unique children | Physician, NP | POHS | Medicaid policies ↑ young children's receipt of POHS and at higher rates in states that required POHS training. These results suggest that oral health training for nondental practitioners is a key component of policy success. |
| Kressin et al.<br>(2009):USA | To assess the effects of this intervention on provider ECC counseling practices, on children's subsequent development of ECC | Multifaceted practice-based intervention | 6 months to 5 years<br>1045 children | Pediatrician, NP, Nurse | Education | Provider knowledge about ECC ↑ after the intervention training. Providers used more counseling strategies. Children at the intervention site had a 77% reduction in risk for developing ECC at follow up. |
| Neumann et al.<br>(2011):Australia | To evaluate the effectiveness of a community-based intervention to improve the oral health of children in non-fluoridated rural Victoria, Australia | Quasi-experimental longitudinal study | 12- < 24 months<br>1013 infant | Maternal and Child Health Nurse (MCHN) | Education | Oral health promotion intervention delivered via local MCHNs promoting early exposure to fluoride may be successful in reducing caries in the 2<sup>nd</sup> year of life but less so in older children when participants have less contact with MCHNs. |
| Long et al.<br>(2012):USA | This study identifies pediatrician-assessed risk factors for ECC and their association with the need for a dentist's evaluation | Cross sectional study | < 3years | Physician | CRA | Caries risk was the strongest predictor of the need for an evaluation. Few patients (6.3%) were classified as high risk. Low referral rates for children with disease and prior to disease onset but at elevated risk, indicate interventions are needed to help improve the dental referral rates of physicians. |
| Herndon et al.<br>(2015):USA | To examine receipt of early childhood caries preventive services (ECCPS) in two states' Medicaid programs before and after the implementation of reimbursement to medical primary care providers (M-PCPs) | - | 6-42 months | Medical<br>Primary Care<br>Provider | PHOS | Reimbursement to M-PCPs was associated with a ↑ likelihood of ECCPS receipt in general and FV application. Reimbursement to M-PCPs can ↑ access to ECCPS. However, ECCPS receipt continues to fall short of recommended care, presenting opportunities for performance improvement. |
| Rolnick et al.<br>(2015):USA | To examine how an intervention to apply FV in a primary health setting to all young, low-income children was implemented and sustained and 2. To assess the feasibility of tracking medical care utilization in this population | - | >6years<br>12,067<br>children | Physician,<br>Pediatrician,<br>NP,<br>Physician<br>Assistant | FV | Of 12,067 children, 85% received FV. Differences were found by age (youngest had highest rates). Small differences by race (81%-88%, highest in Blacks). |
| Veschusio et al.<br>(2016):USA | To examine the effectiveness of South Carolina's Medicaid FV policy on children's receipt of FV from both medical and dental providers | Retrospective | 12-27<br>months<br>52,841<br>children | Physician | FV | The FV rates per child-year were 1 percent for physicians and 23 percent for dentists, respectively. The child-year rate for receipt of FV from both a physician and a dentist was less than one-third of one percent. A policy designed to increase access to FV treatments from physicians and dentists for children up to forty-seven months of age was not successful for physicians. |
| Wigen and Wang<br>(2017):Norway | to evaluate established routines for referral of children by primary care nurses to the dental services and to study whether referred children younger than 3 years required contact with dental personnel | Cross sectional study | <3years<br>181 children | Nurse | Referrals | Majority of children referred were <3 years. Frequent reasons for referral of children <3 years were caries or dental plaque. The referral reasons given by nurses correlated partly with findings at dental examination. Using established referral routines, all children referred from WCV were caries risk children who required early contact with dental personnel. |
| Blackburn et al.<br>(2017):USA | To investigate the effectiveness of preventive dental care in reducing caries-related treatment visits among Medicaid enrollees | Retrospective cohort study | < 2years<br>5095 | Oral health providers (i.e., dentists) vs all other providers (i.e., PCPs) | POHS | 5095 children received preventive dental care before their second birthday, including 3878 from dentists and 1217 from PCPs. Compared with matched children without early preventive dental care, children with dentist-delivered preventive dental care more frequently had a subsequent caries-related treatment, higher rate of visits, and greater dental expenditures. |
| Silk et al.<br>(2018):USA | To implement and study the effect of improving pediatric oral health by training primary care practices and training programs. | - | - | - | POHS | 415 practices across six states were trained, this resulted in 136,963 POHS. Thirty-five of 52 health education programs established pediatric oral health curricula. The average cost of recruitment, training, and follow-up for an office or an educational program was approximately \$1,000/site. |
| Okah et al.<br>(2018):USA | To incorporate OHRAs, including documentation of the oral screening examination, into well-child visits for patients aged 12 to 47 months to drive (1) improved rates of preventive FV application and (2) improved dental referrals for children at high risk for caries | Clinic-based quality improvement study | 12 to 47 months<br>6100 | Pediatrician, NP, Pediatric residents | Screening, FV, Referrals | After multiple Plan-Do-Study-Act cycles, documentation of OHRAs and oral screening examinations improved to 45% and 73%, respectively. The primary outcome measure, FV rates, improved to 86%. Referral of high-risk patients to a dentist improved to 54%. |
| Gnaedinger<br>(2018):USA | The purpose of this QI project was to demonstrate cost-effective implementation of a FV application program at a medium-sized, rural pediatric practice | Cross sectional study | The sample comprised patients aged 9, 18, 24, and 30 months<br>48 children | Pediatrician, Nurse | CRA, FV | 56% of the patients received FV at their WCCs. Economical, FV application program can be readily implemented by nurse practitioners and nurses in pediatric practices where children have inadequate dental care. 20% of children were low risk, 45% medium risk, and 35% high risk. |
| Sibley (2018):USA | To analyze barriers to the implementation of a FV program with a specific focus on cost and reimbursement, as well as assess the costs and benefits of such a program in a pediatric primary care office located in an east central Florida county | Cost benefit analysis | 6 months to 2 years<br>630 | Pediatric primary care office | FV | The data from this cost benefit analysis show a positive financial benefit as an incentive to implement a FV program in this primary care pediatric office and serve as a solid foundation for a future quality improvement project to implement such a program. |
| Dahlberg et al.<br>(2019):USA | To determine if the application of FV to children 5 years and under was acceptable and practical for health care providers in a | Non-randomized experimental study | 0-5 years<br>94 | Family physician, NP | FV | Total direct variable cost of providing FV was \$4.35 per procedure there was an increase from 0% to 9.57% of FV application by providers. Barriers, Potential barriers were lack of proper |
|  | rural primary care office. |  |  |  |  | supplies, lack of adequate support staff, and lack of additional financial compensation for providers. |
| Geiger et al.<br>(2019):USA | To examine a child's odds of receiving POHS in a medical office by county rurality. | Retrospective | < 6 years<br>6,275,456 | Medical providers | POHS | OHS in medical offices were received by 7.8% of children. Rates of POHS in medical offices were higher in metropolitan (metro) counties (8.4%) than nonmetro adjacent to metro (5.8%) and nonmetro not adjacent to metro (4.3%). |
| Kim et al.<br>(2020):USA | to (1) assess use and reimbursement of Current Dental Terminology (CDT) D1206 and Current Procedural Terminology (CPT) 99188 codes, which are the billing codes for FV application; (2) determine when and by whom each FV code was used; and (3) summarize the associated clinical notes | Retrospective | 6- 68 months<br>39 children | Physician, NP, Physician assistant | FV | During the 10-year time period, CDT D1206 was used 5x and CPT 99188 was used 35x. FV was applied exclusively during WCVs. Only pediatricians applied FV in this setting. FV application has been underutilized in this Midwestern tertiary teaching hospital and its affiliated clinics. Advocacy is needed for successful implementation of FV application at WCVs. |
| Johnson and French<br>(2020):USA | The authors sought to utilize quality improvement (QI) methods to increase FV application to reach 85% of eligible well checks at the clinic | Cross sectional study | 6 months to 5 years<br>603 children | Pediatric and psychiatric residents | FV | ↑FV application to 77.7% after cycle 1, and 74% in cycle 2. Documentation increased after prompt. Brief educational interventions may result in ↑ use of FV in resident-based clinics. Task based prompts or stop measures in EMR templates can improve documentation, which can inform efforts to improve FV application. |
| Roth et al.<br>(2020):USA | Through a resident-led quality improvement (QI) project, we aimed to provide | - | 1 year to 5 years<br>323 children | Residents | FV | The resident-led QI project ↑ rates of FV application, dental referrals, and dental visits while meeting ACGME guidelines |
|  | FV to 50% of patients ages 1 through 5 who did not have a dental visit in the preceding 6 months or receive FV elsewhere in the past month. |  |  |  |  | for experiential learning in QI. By adapting to state-specific guidelines and workflows of each clinic, this QI project could be nationally reproduced to improve adherence to AAP and USPSTF guidelines. |
| Zea and Henshaw (2022):USA | The study assessed the level of medical-dental integration achieved by the pilot and maintained over 2 years after program implementation | Retrospective | 72 months of age or younger<br>1,456 children | Pediatrician | Screening, FV | The proportion of WCVs during which pediatricians integrated oral health preventive measures ↑ by 25% to 50% from baseline (2014) to the end of the pilot (2016) and by at least 5% from 2016 to 2018. |
| Melgar et al. (2024):Peru | To explore the association between CRED and oral health services utilization (OHSU), throughout the heterogeneous Peruvian territory | Cross sectional study | 12 to 59 months<br>15,836 children | Primary health care professionals | Referral | Integrating oral health into Peruvian Child PHC seems to be a promising public health intervention to ↑ children's OHSU. |
| Verlinden et al. (2024):Netherlands | To evaluate whether active or passive referral by a well-childcare (WCC) physician of babies for a first preventive dental visit leads to earlier initiation of dental care. | Quasi-experimental study | children ages 4-11 months<br>1347 children | Physicians | Referrals | Of the active referral intervention group, 59.3% had their first preventive dental visit in their first year, for the passive referral group, 46.9%. Referral of babies by WCC for their first preventive dental visit led to earlier initiation of dental care. An active referral had a larger effect than passive referral |
| Ahmed et al.<br>(2021):USA | To longitudinally assess the association between age at first oral health examination and provider type at first oral health examination on dental treatment for children under 6 y of age | Retrospective | 6 to < 6<br>years<br>2,419,026<br>Children | Family<br>physician;<br>General<br>dentists,<br>pediatric<br>dentists | Screening | Dental caries for children whose first oral health examination at 4yrs was 5x higher than those before 1 y of age. Dental caries for children seen by pediatric dentists and physicians was ↑ than those seen by a general dentist. Study highlights the importance of first oral health examination no later than 12 months of age and referrals from physicians to prevent the need for invasive treatment. |
| Braun et al.<br>(2015):USA | To understand the impact of Colorado's interprofessional OHE program on health care professionals and practice behaviors around the provision of OHPS to children, and to identify factors that facilitated or created barriers to its diffusion | Quasi-experimental<br>OHP<br>intervention | children<br><5years | Sampling<br>medical,<br>dental, and<br>nontraditional | Education,<br>Screening,<br>CRA, FV | From 2009 to 2012, the proportion of young, low-income children receiving OHPS from a medical professional ↑16-fold. Many practices had initiated practice-level changes to support program implementation. Factors facilitating program diffusion -quality materials, community need, and reimbursement; barriers included lack of time to provide services, resources to purchase supplies, and referral dentists. |
| Gracner et al.<br>(2023):USA | To examine monthly changes in FV applications among pediatric clinicians following the ACA mandate | Observational<br>cohort study | 2405<br>clinicians,<br>with 107<br>841<br>clinician-<br>months | Physician,<br>Pediatrician,<br>Clinicians | FV | Premandate, 10.48% of the visits included FV applications. Two years postmandate, the likelihood of applying FV was 13.64% points higher. For clinicians providing FV premandate, the share of visits with FV ↑ by 9.22% points. This ↑ was observed in clinicians who treated children with insurance that was mostly mixed and mostly private, no substantial change with public insurance. |
| Abreu-Placeres et al. (2023):<br>Dominican Republic | To test the effectiveness in the prevention of ECC through an educational intervention program with the use of a printed guide for pediatricians and parents both designed by pediatric dentists | Two-arm RCT | 10 to 12 months<br>309 children | Pediatrician | Education | The RCT showed no effectiveness in caries-progression control. Despite this result, the study managed to identify barriers that do not allow pediatricians from offering parents adequate oral health recommendations. |
| Lukac et al. (2023):USA | To increase the application of FV in children while analyzing the effect of two passive CDS tools—an order set and a note template | Retrospective cohort | 6 months to 5 years<br>3049 well-child visits | Resident physicians and Attending physicians? | FV | The addition of a fluoride order to a “Well Child Check” order set led to a 10.6% ↑ in ordering over physician education alone. The insertion of fluoride-specific text to drop-down lists in clinical notes led to a 6.2% ↑. The targeted use of order sets and note templates was positively associated with the ordering of topical FL by physicians. Added revenue totaled \$15,084. |
| Danesh et al. (2023):USA | To evaluate child-level dental utilization and expenditure outcomes based on if and where children received FV at quality improvement (QI) medical practices, at non-QI medical practices, at dental practices, or those who never received FV from any practice. | Retrospective claims-based analysis cohort study. | 1-5 years<br>98,001 children | Participating practices<br>Medical and dental | FV | Children who received FV at QI medical practices had a significantly higher incidence rate of preventive dental visits than children who received FV at dental practices or non-QI medical practices. Children who received FV only at dental practices were significantly more likely to have a dental GA visit than children who received FV at QI medical practices. |
| Giles et al.<br>(2022):USA | To conduct an early-phase feasibility study of an oral health intervention, Health visitors delivering Advice on Britain on Infant Toothbrushing, delivered by Health Visitors to parents of children aged 9–12 months old. | A mixed-methods, early-phase, non-controlled, feasibility study. | 9-12 months<br>35 children | Health visitors | Tooth brushing | Total compliance with toothbrushing guidelines at baseline was low (30%) but significantly improved and was maintained 3 months after the intervention (68%). Plaque scores improved post intervention and participants found video recording of toothbrushing acceptable. Dietary habits remained largely unchanged. |
| Ko et al.<br>(2022):USA | To examine physician and dentist FL prescription patterns and identify the factors associated with FL prescriptions for Medicaid-enrolled children. | Secondary analysis of administrative Medicaid enrollment and claims data | 200,169 children | Physician, Pediatrician, NP; Physician assistant | FV | Of 200,169 Medicaid-enrolled children, 6.7% received FL prescriptions. A larger proportion of children aged <6 years received FL. Physicians were >3 times as likely to prescribe FL as dentists. Children with a history of FL prescriptions and any restorative dental treatment were significantly more likely to receive a FL prescription, whereas children living in areas with water fluoridation were significantly less likely. Physicians play an important role in prescribing fluoride to Medicaid-enrolled children, especially those at increased dental caries risk. |
| Brännemo et al.<br>(2021):Sweden | To evaluate oral health outcomes and early oral health promotion of children in a Swedish, parental support program conducted in a collaboration between Child Health Services and Social Services. | Non-randomized experimental study | 0 to 15 months<br>recruitment follow-up at 18 and 36 months<br>201 children<br>101 intervention- | Child Health Care nurse and a parental advisor from Social Services | Education | ↓ Caries prevalence and tooth brushing habits more consistent in the intervention group. Parents in the intervention group introduced tooth brushing twice daily more often when their child was 18 months. The proportion of children with cavitated caries lesions (ICDAS 3-6) at 18 months was significantly ↓ in the intervention group. The extended |
|  |  |  | 100 in<br>control |  |  | postnatal home visiting program had a positive impact on oral health. |
| Turton et al.<br>(2021):Asia-Cambodia | To critically review the feasibility of the Cambodia Smile intervention by considering clinical outcomes, acceptability and stakeholder perceptions | Mixed method | 6 to 24<br>months<br>intervention<br>group<br>392 children | nurses and<br>midwives | Education, FV | OHE and FV interventions provided by NDPCPs were feasible and acceptable for stakeholders in a Cambodian setting. The intervention group had lower ECC experience and better OHRQoL at 2 years of age. Surveyed parents had favorable views of the FV placement by medical professionals. |
| Kranz et al.<br>(2020):USA | To test the hypothesis that widespread adoption of state Medicaid policies supporting medical POHS may have unintentionally reduced dental visits | Cross sectional | 6mo to <6y<br>45.1million<br>child-years | Medical and<br>Dental<br>preventive<br>POHS | POHS | The study failed to find evidence that medical POHS replaced dental visits for young children enrolled in Medicaid and, offers evidence that increased medical POHS was associated with increased utilization of dental care. Given lower-than-desired rates of dental visits for this population, delivery of medical POHS should be expanded. |
| Kranz et al.<br>(2014a):USA | To examine the distribution of dental and medical Medicaid providers of pediatric oral health services throughout North Carolina to determine if these services have improved access to care for Medicaid enrollees <3 yrs old. | Retrospective | < 3 years | Physician | POHS | The study underscores how physician-based POHS can help increase the geographic availability of oral health services for young Medicaid enrollees, enabling the delivery of oral health services in areas not served by a dental provider. |
| Kranz et al.<br>(2015):USA | To evaluate the impact of comprehensive POHS, provided in medical offices by NDPCPs, on the dental caries experience of children enrolled in North Carolina's (NC's) Medicaid program | Retrospective | 29 173 kindergarten students | Non dental providers | POHS | POHS provided by NDPCPs in medical settings were associated with a ↓ in caries experience in young children but were not associated with improvement in subsequent use of treatment services in dental settings. |
| Turner et al.<br>(2010):Scotland | The study describes monitoring arrangements and summarizes data covering the period 2006-2009 of the Childsmile program. | Observational |  | Health Visitors | POHS | By mid-2009, around 28,000 infants in deprived areas of the West of Scotland had been given CRA by Health Visitors; 14,000 were enrolled with 142 Childsmile practices or clinics; and over 10,000 had begun making practice visits. The Childsmile Nursery and School programs had provided 28,000 FV treatments to nursery and primary school children. |

WCV- Well child visit, FV- Fluoride varnish, NP- Nurse practitioner, MA- Medical assistant, FVT- Fluoride varnish therapy, IMB- into the Mouths of Babies, PCP-primary care provider, dmf- decayed missing filled (cumulative score of the number of decayed, missing due to caries and filled primary teeth), ECOH- early childhood oral health

### Target Population and Settings

All studies focused on children under six years of age, with a majority targeting infants and toddlers under three years (Abreu-Placeres et al., 2023; Brännemo et al., 2021; Pahel et al., 2010; Turton et al., 2021). Interventions were commonly implemented in primary care clinics, community health centers, well-child visit settings, and maternal-child health programs. Many of the studies specifically targeted Medicaid-enrolled populations, underserved communities, and Indigenous or rural populations. Additional settings included home visitation programs, school-based initiatives, and community outreach through child health nurses and health visitors.

### Non-Dental Provider Types and Training

NDPCPs engaged in oral health service delivery included physicians (pediatricians and family physicians), nurse practitioners (NPs), registered nurses, physician assistants, medical assistants, health visitors, dietitians, and trainees. Physicians (n = 33) and nurse practitioners (n = 14) were most frequently involved (Figure 3). Interprofessional approaches were often described, with teams that integrated nurses, community health workers, and dietitians.

**Figure 3:**
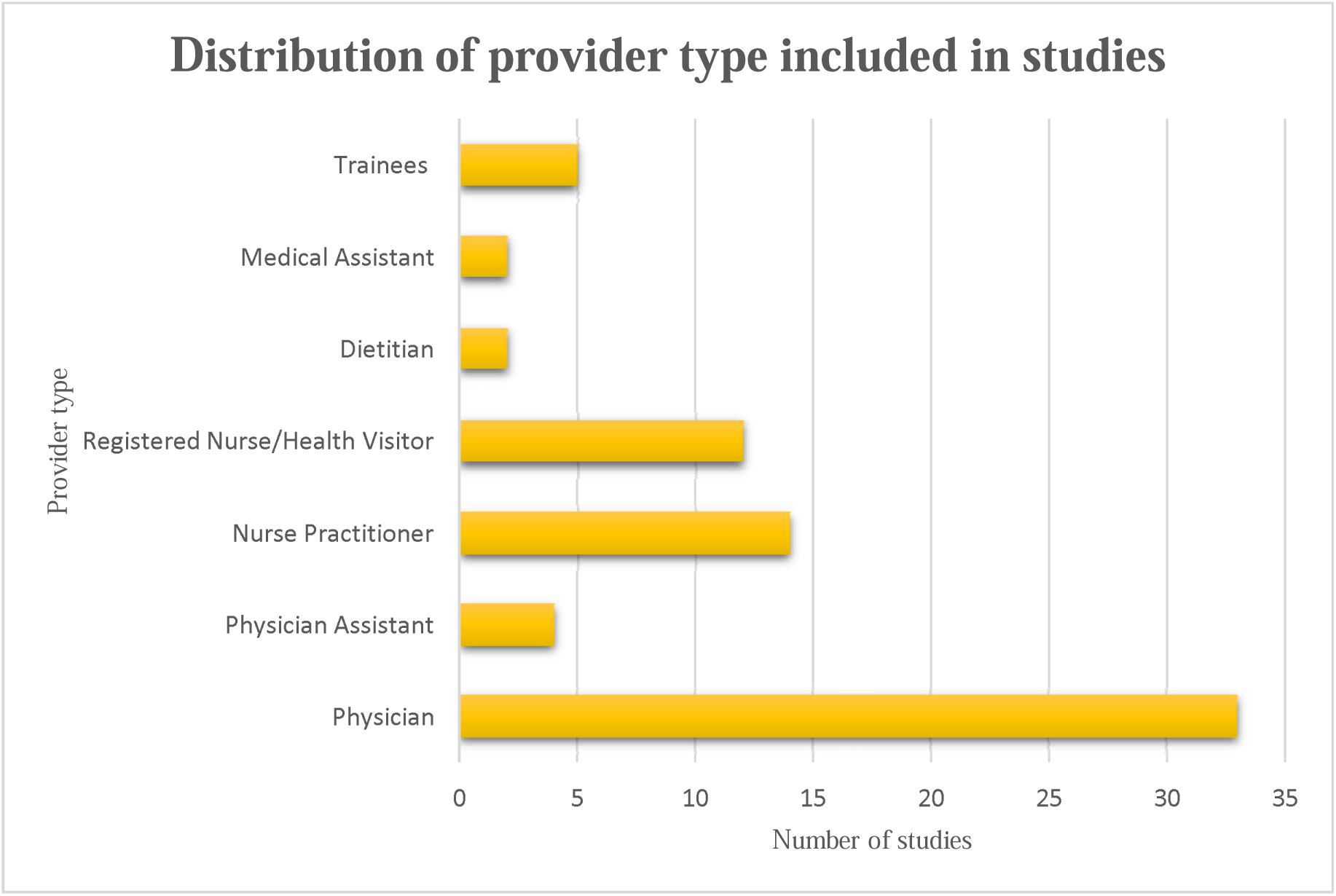
Distribution of provider type included in studies.

Training programs were a key enabler of successful implementation. Training formats ranged from brief online modules and recorded videos to multi-hour workshops and structured curricula like the “Smiles for Life” program (Cheng et al., 2019; Dahlberg et al., 2019; McCulley et al., 2022; Murphy & Larsson, 2017; Silk et al., 2018). Programs included didactic instruction, role-play, clinical photos, (Biordi et al., 2015; Kressin et al., 2009; Long et al., 2012) visual aids such as posters, brochures and handouts were also used to facilitate training (Braun et al., 2015; Neumann et al., 2011) and competency assessments (Taylor et al., 2014). Training was context-specific, as seen in Turton et al. (2021), where community nurses and midwives received an initial 8-hour training followed by periodic refreshers. Tailoring to practice setting was also important: for example, (Turner et al., 2010) trained health visitors for home-based CRA and referral, while Maher et al. (2012) used motivational interviewing techniques and culturally adapted educational tools in Australia’s Aboriginal communities.

### Interventions Delivered by Non-Dental Providers

NDPCPs provided a variety of oral health interventions, including oral health education, CRA, fluoride varnish application, dental referrals, and EMR-integrated care. Their delivery varied based on the clinical setting, provider type, population served, and training received.

### Oral Health Education

Educational interventions were widely reported and typically delivered during well-child visits or home/community encounters. These interventions aimed to improve caregiver knowledge and promote early preventive behaviors such as daily brushing, fluoride toothpaste use, and avoidance of sugary snacks. Tools included pictorial flipcharts (Cheng et al., 2019), brochures (Kressin et al., 2009), posters (Maher et al., 2012), handouts (Gnaedinger, 2018), quiz and factsheets (Berger et al., 2014). Educational content was often culturally tailored and age-appropriate (Roth et al., 2020), with messages integrated into existing child health records or reinforced through in-room reminders (Okah et al., 2018). Community-based delivery was observed in home visits (Brännemo et al., 2021; Giles et al., 2022; Turner et al., 2010) and through outreach services distributing toothbrushes and toothpaste (Turton et al., 2021).

### Caries Risk Assessment

CRA was increasingly adopted by NDPCPs as a core preventive strategy, often facilitated through structured tools or checklists. These tools were either adapted from existing dental frameworks or specifically designed for medical settings. For instance, multiple studies used variations of the AAP Oral Health Risk Assessment Tool (Jackson, 2015; McCulley et al., 2022; Okah et al., 2018), or the CAMBRA model (Biordi et al., 2015). Berger et al. (2014) implemented the AAPD Caries Risk Assessment Tool, while Cheng et al. (2019) employed the Nursing Caries Assessment Tool (N-CAT), a self-administered tool completed by families within 5 to 10 minutes. Gnaedinger (2018) also described use of a dental CRA tool that helped guide provider decision.

CRA was often integrated into routine well-child or primary care visits. Braun et al. (2017) described CRA performed by physicians, NPs, and PAs as part of a broader *Cavity Free at Three* program. Similarly, Pahel et al. (2010) reported the use of structured encounter forms to document CRA alongside POHS. In some interventions, CRA was supported by EMR modifications and prompts, facilitating documentation and follow-up. For example, Okah et al. (2018) included EMR templates that prompted documentation of CRA, fluoride varnish, and referrals, while Sudhanthar et al. (2019) embedded automatic CRA-related reminders and fluoride orders into the EMR system.

Overall, CRA served as a pivotal step in identifying children at elevated risk for dental disease, enabling providers to deliver tailored anticipatory guidance, apply fluoride varnish, and refer to dental services as needed.

### Fluoride Varnish Application

Fluoride varnish application was the most reported clinical intervention delivered by NDPCPs (Table 2). Providers applied fluoride varnish during well-child visits, WIC appointments, and community outreach activities. Studies overwhelmingly reported that fluoride varnish application was feasible, acceptable, and effective in reducing ECC incidence when applied consistently.

**Table 2:**
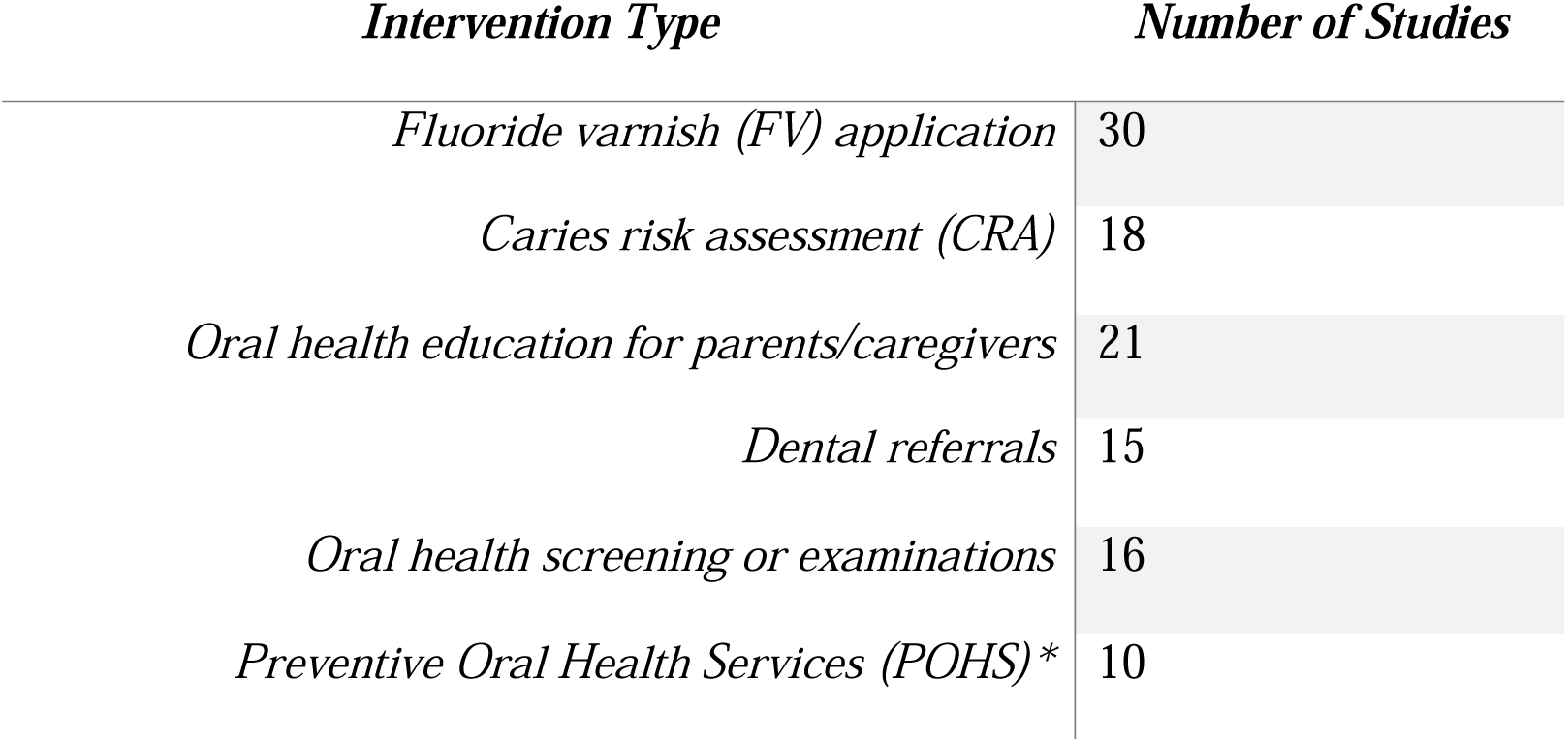
Intervention type and number of studies.

Programs such as *Into the Mouths of Babes* in North Carolina (Pahel et al., 2011; Rozier et al., 2010) demonstrated that multiple fluoride varnish applications delivered by physicians significantly reduced caries-related treatment needs. Braun et al. (2017) found that receiving at least four fluoride varnish applications by age three was associated with a significant reduction in ECC.

Several studies reported increases in fluoride varnish uptake following provider training and implementation of quality improvement initiatives (Cheng et al., 2019; Murphy & Larsson, 2017; Sudhanthar et al., 2019). For example, in a resident-led intervention, Roth et al. observed a substantial increase in fluoride varnish application and dental referrals (Roth et al., 2020). Policy and reimbursement incentives also played a role; Okunseri et al. (2009), Okunseri et al. (2010), Herndon et al. (2015), and Kranz et al. (2022) found that Medicaid reimbursement policies increased fluoride varnish claims by medical providers.

Barriers to implementation, such as time constraints, lack of supplies, and billing challenges, were noted by providers in some studies (Dahlberg et al., 2019; Kim et al., 2020). Nevertheless, the broad adoption of fluoride varnish by NDPCPs suggests strong potential for sustainability and scalability.

## Dental Referrals

Dental referrals by NDPCPs were instrumental in ensuring continuity of care for children identified as being at risk for ECC. Referral decisions were typically based on CRA findings, screening outcomes, or parental concern.

Several studies reported improvements in referral practices following training or the introduction of structured tools. McCulley et al. (2022) observed a 100% improvement in dental referrals after integrating CRA into well-child visits. Okah et al. (2018) improved referral rates to 54% for high-risk patients following EMR and workflow enhancements. Jackson (2015) similarly demonstrated increased identification and referral of high-risk children with the use of a structured risk assessment tool.

In community-based models, referral pathways were integrated into routine care. Turner et al. (2010) described how Scottish health visitors used CRA to refer children to Childsmile dental services, while Wigen and Wang (2017) found that referrals from well-baby clinics effectively identified high-risk children. In Peru, Melgar et al. (2024) highlighted the effectiveness of embedding referrals into the CRED program to improve utilization of dental services.

Importantly, the mode of referral impacted its effectiveness. Verlinden et al. (2024) found that active referral, where the provider directly facilitated the dental visit was significantly more effective than passive referral methods in promoting early dental visits.

Despite these successes, low baseline referral rates and under-identification of at-risk children in some settings underscored the need for ongoing provider support and system-level reinforcement (Long et al., 2012; Patton & Severe, 2020).

### Integration into EMRs

Integration of oral health interventions into EMRs was a critical enabler for sustainable service delivery. EMR prompts, structured documentation templates, and decision support tools helped embed oral health practices into routine care and facilitated consistent follow-up. Okah et al. (2018) implemented a multifaceted EMR strategy that included prompts for CRA, fluoride varnish application, and referrals, leading to improved documentation and service uptake. Similarly, Lukac et al. (2023) found that adding fluoride varnish –specific order sets and note templates in the EMR significantly increased the likelihood of fluoride varnish being ordered and applied.

EMRs were also used to support quality improvement efforts. Johnson and French (2020) utilized task-based prompts to improve fluoride varnish documentation and application rates among pediatric residents, while Roth et al. (2020) sent monthly reminders and personalized performance feedback to improve provider adherence. McCulley et al. (2022) and Gnaedinger (2018) highlighted how EMR integration enhanced the documentation of CRA and preventive care. These findings underscore the value of health information systems in promoting oral health equity by standardizing care delivery and closing service gaps.

Many interventions were embedded into existing clinical workflows. Jackson (2015) and Okah et al. (2018) included EMR prompts and clinical decision tools to standardize oral health documentation and support application of fluoride varnish. Other studies leveraged interprofessional collaboration, with dental professionals often serving as trainers for medical providers (Berger et al., 2014; Cheng et al., 2019; Okah et al., 2018).

Overall, the evidence indicates that targeted, practice-integrated training particularly when reinforced by EMR prompts and supported by dental professionals can build capacity among NDPCPs to deliver effective oral health risk assessment and preventive care in primary care settings.

## Discussion

This scoping review identified and synthesized evidence from 54 studies examining the role of NDPCPs in delivering CRA and POHS to children under six years of age. The findings highlight growing international efforts to integrate oral health into primary care settings and underscore the feasibility, acceptability, and potential effectiveness of this interprofessional approach in ECC prevention.

### Expanding Roles of Non-Dental Providers in Oral Health

Across the included studies, physicians, nurse practitioners, registered nurses, physician assistants, health visitors, and allied professionals such as dietitians played key roles in delivering POHS including oral health education, CRA, fluoride varnish application, and dental referrals. The involvement of NDPCPs addresses oral health workforce gaps and increases access for vulnerable populations, especially in rural, Indigenous, and Medicaid-enrolled communities (Cheng et al., 2019; Kranz et al., 2014a; Turner et al., 2010).

These findings support earlier assertions that NDPCPs are well positioned to deliver oral health interventions during routine child health visits (McCulley et al., 2022; Rozier et al., 2010).

Studies also demonstrate that targeted oral health training builds provider competence and improves service delivery (Braun et al., 2017; Silk et al., 2018).

### Training Improves Confidence, Uptake, and Outcomes

The review confirms that NDPCP training is foundational to successful integration of POHS. Structured educational interventions ranging from brief modules to full-day workshops significantly improved knowledge, CRA documentation, fluoride varnish application, and referral practices (Biordi et al., 2015; Cheng et al., 2019; Maher et al., 2012). The “Smiles for Life” curriculum and role-play techniques enhanced practical skills and provider confidence (Kressin et al., 2009; Silk et al., 2018).

Competency-based approaches further ensured skill retention and performance, with some programs embedding training into WIC services (Taylor et al., 2014) or offering refresher sessions for sustained impact (Turton et al., 2021).

However, persistent barriers such as limited time, insufficient supplies, and unclear billing mechanisms were noted (Braun et al., 2015; Dahlberg et al., 2019). These challenges highlight the importance of system-level supports to complement provider training and encourage sustained practice change. Similar barriers have been reported among non-dental primary care providers in Manitoba, Canada, where limited appointment time, inadequate supplies, and billing ambiguity constrained the integration of CRA and preventive oral health services into primary care

### Effectiveness of Interventions in Preventing ECC

NDPCPs delivered a range of effective interventions that contributed to improved oral health behaviors and outcomes. Oral health education, especially when culturally tailored, promoted early toothbrushing, fluoride toothpaste use, and dietary changes (Brännemo et al., 2021; Giles et al., 2022; Maher et al., 2012). Some studies integrated education into existing health records or provided take-home materials to reinforce learning (Cheng et al., 2019; Murphy & Larsson, 2017).

CRA emerged as a pivotal practice for risk identification and guided follow-up actions such as fluoride varnish and referrals. Tools like the AAP Oral Health Risk Assessment and Nursing Caries Assessment Tool facilitated structured screening (Cheng et al., 2019; McCulley et al., 2022). When CRA was embedded in clinical workflows, providers were better able to tailor preventive services to high-risk children (Jackson, 2015; Okah et al., 2018).

Fluoride varnish application, reported in most studies, was consistently linked with reductions in ECC incidence when delivered at recommended intervals (Braun et al., 2017; Rozier et al., 2010). Multiple studies reported significant increases in fluoride varnish rates following quality improvement interventions and Medicaid reimbursement policy changes (Herndon et al., 2015; Kranz et al., 2022; Pahel et al., 2011).

### Referral Practices and Early Dental Visits

Referrals by NDPCPs played a critical role in bridging children to dental care, particularly those at high risk. Studies such as McCulley et al. (2022) and Okah et al. (2018) demonstrated improved referral rates following EMR enhancements and provider training. Programs that incorporated CRA into home visits or maternal-child programs such as Childsmile in Scotland (Turner et al., 2010) or the Peruvian CRED model (Melgar et al., 2024) showed promise in increasing early dental visits.

Importantly, the method of referral influenced outcomes. Verlinden et al. (2024) found that active referrals, where providers directly facilitated the dental appointment, were significantly more effective than passive strategies. However, persistently low referral rates in some studies (Long et al., 2012; Patton & Severe, 2020) indicate ongoing challenges in linking at-risk children to appropriate dental care, especially in underserved areas.

### Integration into EMRs Supports Sustainability

EMR integration emerged as a key enabler for sustained oral health service delivery. Studies consistently found that EMR prompts, templates, and decision support tools improved documentation and uptake of CRA, fluoride varnish, and referrals (Lukac et al., 2023; Okah et al., 2018). Quality improvement projects further leveraged EMR systems to provide feedback and monitor provider performance (Johnson & French, 2020; Roth et al., 2020).

Embedding oral health into EMRs also facilitated adherence to guidelines and allowed providers to manage time more efficiently. This approach supports the broader call for health system reforms that institutionalize POHS within routine primary care (Kranz et al., 2015; Zea & Henshaw, 2022). While there is limited published data on EMR integration of oral health in Canadian primary care settings, emerging implementation efforts in Manitoba highlight provider interest in digital tools to support CRA documentation and referral pathways (Olatosi et al., 2025b). As such, integrating CRA and POHS into EMRs should be considered a priority in Canada to enable workflow efficiency, documentation fidelity, and long-term sustainability.

### Implications for Policy and Practice

The findings of this review align with recommendations from organizations such as the United States Preventive Services Task Force (USPSTF), which endorses fluoride varnish application for all children under five and emphasizes the role of medical providers in ECC prevention (Davidson et al., 2021). Evidence from this review also supports policy strategies such as Medicaid reimbursement for medical providers, mandatory oral health training, and the use of EMR systems to scale up delivery of POHS in medical settings (Kranz et al., 2020; Okunseri et al., 2009).

Furthermore, interprofessional collaboration, stakeholder engagement, and culturally sensitive approaches were integral to successful implementation. Future efforts should continue to prioritize training, streamline referral pathways, strengthen EMR infrastructure, and address systemic barriers that limit access and uptake, particularly in rural and marginalized communities.

Notably, the vast majority of studies included in this review originated from the United States, with a small number from other countries such as Australia, Sweden, Scotland, and Peru. These findings suggest that the integration of oral health into primary care by NDPCPs is a growing global trend, particularly in settings where health systems support interprofessional collaboration and reimbursement for preventive services. However, no studies meeting the inclusion criteria were identified from Canada. This gap is significant given the country’s universal healthcare aspirations and increasing awareness of the oral-systemic health connection.

The absence of Canadian studies meeting our inclusion criteria may reflect systemic gaps, including the lack of national policies, provider training requirements, and reimbursement frameworks to support oral health service delivery by NDPCPs. Nevertheless, several emerging initiatives in Canada are beginning to address these challenges. For instance, ElSalhy et al. (2019) explored pediatric residents’ perspectives on the feasibility of integrating preventive dental care into a general pediatric outreach clinic serving a First Nations community, highlighting interest and potential in expanding provider roles. In Ontario, Da Silva et al. (2020) examined the views of key stakeholders on incorporating fluoride varnish application into routine primary care, identifying both enablers and barriers to implementation. More recently, Hachey et al. (2020) advocated for interprofessional collaboration and the integration of core oral health services such as screening, parental counseling, fluoride varnish application, and referrals into routine primary care for Canadian children.

Notably, in December 2019, the Public Health Agency of Canada supported the development of a novel Canadian Caries Risk Assessment tool for preschool-aged children (Schroth et al., 2021). Designed for use by NDPCPs, this tool facilitates the identification of caries risk, delivery of anticipatory guidance, application of fluoride varnish, and referral of high-risk children to dental providers. These foundational efforts signal growing recognition of the need to embed oral health into early childhood care and reinforce the case for coordinated policy, funding, and training strategies to promote oral health equity, particularly for Indigenous and rural populations.

## Conclusion

This scoping review provides a comprehensive synthesis of the growing body of evidence supporting the integration of CRA and POHS into primary care for children under six years of age. NDPCPs, including physicians, nurses, and allied health professionals, are increasingly recognized as valuable contributors to ECC prevention. Through training, interprofessional collaboration, and supportive system-level structures such as EMR integration and reimbursement policies, NDPCPs have successfully delivered CRA, fluoride varnish, and oral health education in a variety of settings. These efforts have demonstrated positive impacts on early dental visits, ECC prevention, and caregiver engagement particularly in underserved, rural, and Indigenous populations. The findings of this review reinforce the importance of embedding oral health into early childhood preventive care and offer practical insights to guide program development, policy implementation, and future research.

## Limitations

Several limitations must be considered when interpreting the findings of this review. First, although the review included a diverse range of study designs and international contexts, most studies were conducted in the United States, potentially limiting generalizability to other healthcare systems. Second, many studies used retrospective designs and administrative data, which may underreport outcomes such as fluoride varnish application or referrals due to incomplete documentation. Third, variability in study quality and intervention fidelity was noted, with some lacking detailed descriptions of training content, implementation processes, or outcomes measured. Additionally, while this review captured a wide range of interventions, few studies explicitly assessed long-term sustainability or the cost-effectiveness of NDPCP-led oral health programs. Finally, this review did not conduct a formal quality appraisal of included studies, consistent with scoping review methodology, but this limits the ability to assess the strength of the evidence base.

## Future Directions

Building on the promising evidence identified, future research should explore the long-term impact and sustainability of oral health integration into primary care, particularly in non-U.S. contexts and in collaboration with Indigenous communities. Mixed-method and implementation science approaches can help uncover the mechanisms, facilitators, and barriers to scale-up across different health system levels. Economic evaluations would also be valuable in understanding cost-effectiveness, particularly for training programs, EMR integration, and reimbursement models. Additionally, more studies are needed that center the perspectives of caregivers, community members, and front-line providers to ensure interventions are culturally responsive and contextually appropriate. Policymakers and health system leaders should consider advancing oral health equity by supporting interprofessional education, enhancing access to EMR infrastructure, and aligning incentives to promote the routine inclusion of oral health services in well-childcare.

**Appendix 1:**
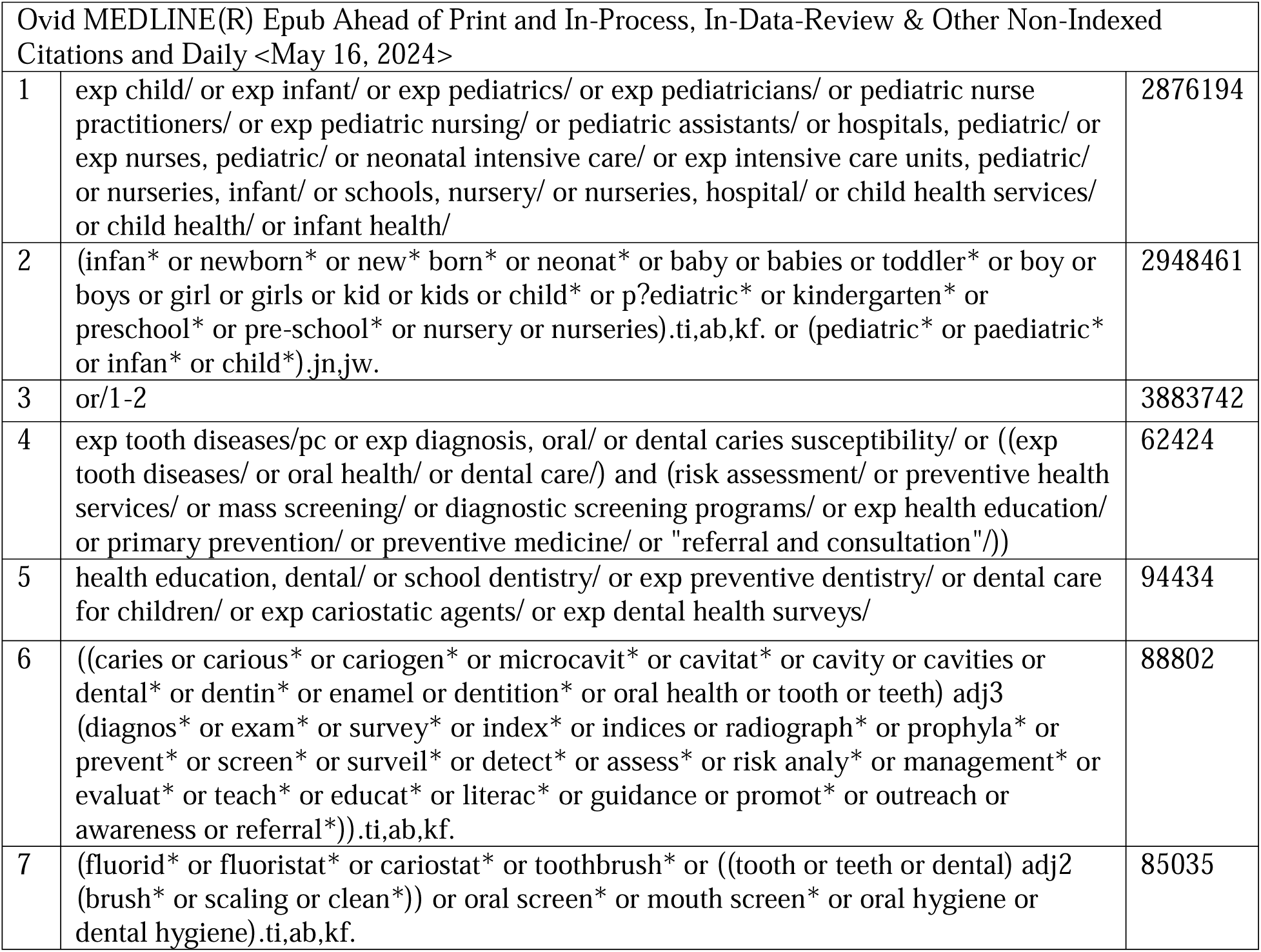
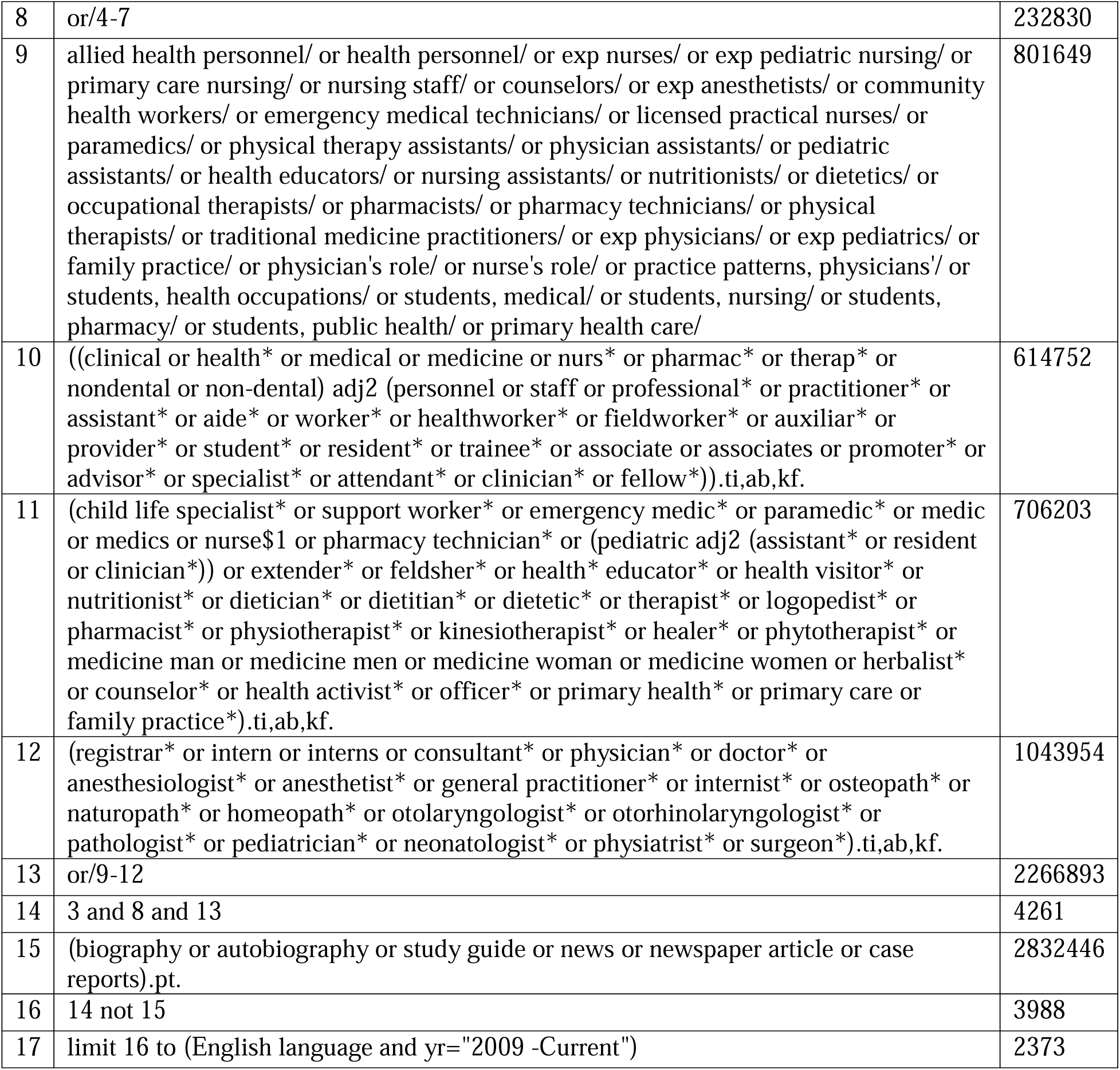
Search strategy with results from Ovid MEDLINE(R) database.

## Author Contributions

OOO substantially contributed to the conceptualization and identification of research questions, study selection (title and abstract screening), data charting and collation, summarizing and reporting of results, and drafting of the manuscript. RJS contributed to the conception, study design, data analysis, and critically revised the manuscript for important intellectual content. JV, CY and MO contributed to data acquisition, analysis, and critically revised the manuscript for important intellectual content. Nicole Askin contributed to study design and data acquisition. JS, PC, and JL critically revised the manuscript for important intellectual content.

## Data Availability

All data produced in the present study are available upon reasonable request to the authors

## References

1. Abreu-Placeres, N., Ekstrand, K. R., Garrido, L. E., Bakhshandeh, A., & Martignon, S. (2023). An interdisciplinary intervention program to prevent early childhood caries in the Dominican Republic. FRONTIERS IN ORAL HEALTH, 4, Article 1176439. 10.3389/froh.2023.1176439

2. Ahmed, I., McGivern, S., Beymer, M. R., Okunev, I., Tranby, E. P., Frantsve-Hawley, J., Tseng, C. H., & Ramos-Gomez, F. (2021). Age of First Oral Health Examination and Dental Treatment Needs of Medicaid-Enrolled Children. JDR Clin Trans Res, 23800844211057793. 10.1177/23800844211057793

3. Arksey, H., & and O’Malley, L. (2005). Scoping studies: towards a methodological framework. International Journal of Social Research Methodology, 8(1), 19–32. 10.1080/1364557032000119616

4. Berger, C., Bachman, J., Casalone, G. G., Farberman, S., & Fish, A. (2014). An oral health program for children. Nurse Pract, 39(2), 48–53. 10.1097/01.Npr.0000441912.47395.46

5. Biordi, D. L., Heitzer, M., Mundy, E., DiMarco, M., Thacker, S., Taylor, E., Huff, M., Marino, D., & Fitzgerald, K. (2015). Improving access and provision of preventive oral health care for very young, poor, and low-income children through a new interdisciplinary partnership. Am J Public Health, 105 Suppl 2(Suppl 2), e23–29. 10.2105/ajph.2014.302486

6. Blackburn, J., Morrisey, M. A., & Sen, B. (2017). Outcomes Associated With Early Preventive Dental Care Among Medicaid-Enrolled Children in Alabama. JAMA Pediatr, 171(4), 335–341. 10.1001/jamapediatrics.2016.4514

7. Brännemo, I., Dahllöf, G., Cunha Soares, F., & Tsilingaridis, G. (2021). Impact of an extended postnatal home visiting programme on oral health among children in a disadvantaged area of Stockholm, Sweden. Acta Paediatr, 110(1), 230–236. 10.1111/apa.15457

8. Braun, P. A., Racich, K. W., Ling, S. B., Ellison, M. C., Savoie, K., Reiner, L., & Westfall, J. M. (2015). Impact of an interprofessional oral health education program on health care professional and practice behaviors: a RE-AIM analysis. Pediatric Health Med Ther, 6, 101–109. 10.2147/phmt.S79826

9. Braun, P. A., Widmer-Racich, K., Sevick, C., Starzyk, E. J., Mauritson, K., & Hambidge, S. J. (2017). Effectiveness on Early Childhood Caries of an Oral Health Promotion Program for Medical Providers. American Journal of Public Health, 107, S97–S103. 10.2105/AJPH.2017.303817

10. Canada’s Drug Agency. (2016). Peer Review of Electronic Search Strategies: 2015 Guideline Explanation and Elaboration (PRESS E&E).

11. Cheng, J. K., Faniyan, A., Chan Yuen, J., Myers, T., Fleck, M., Burgess, J., Williams, K., Wijeratne, R., Webster, R., Cox, J., & Ng, M. W. (2019). Changes in Oral Health Behaviors Associated With a Nursing Intervention in Primary Care. Glob Pediatr Health, 6, 2333794x19845923. 10.1177/2333794x19845923

12. Colak, H., Dulgergil, C. T., Dalli, M., & Hamidi, M. M. (2013). Early childhood caries update: A review of causes, diagnoses, and treatments. J Nat Sci Biol Med, 4(1), 29–38. 10.4103/0976-9668.107257

13. Da Silva, K., Daniel, I., Singhal, S., Feller, A., & Quiñonez, C. (2020). The Use of Fluoride Varnish in Primary Care in Ontario: A Qualitative Study. J Can Dent Assoc, 86, k6.

14. Dahlberg, D., Hiott, D. B., & Wilson, C. C. (2019). Implementing Pediatric Fluoride Varnish Application in a Rural Primary Care Medical Office: A Feasibility Study. Journal of Pediatric Healthcare, 33(6), 702–710. 10.1016/j.pedhc.2019.06.002

15. Danesh, D. O., Peng, J., Hammersmith, K. J., Gowda, C., Maciejewski, H., Amini, H., Wapner, A. W., & Meyer, B. D. (2023). Impact on Dental Utilization of the Integration of Oral Health in Pediatric Primary Care Through Quality Improvement. Journal of Public Health Management & Practice, 29(2), 186–195. 10.1097/PHH.0000000000001689

16. Davidson, K. W., Barry, M. J., Mangione, C. M., Cabana, M., Caughey, A. B., Davis, E. M., Donahue, K. E., Doubeni, C. A., Kubik, M., Li, L., Ogedegbe, G., Pbert, L., Silverstein, M., Stevermer, J., Tseng, C.-W., & Wong, J. B. (2021). Screening and Interventions to Prevent Dental Caries in Children Younger Than 5 Years: US Preventive Services Task Force Recommendation Statement. JAMA: Journal of the American Medical Association, 326(21), 2172–2178. 10.1001/jama.2021.20007

17. Douglass, A. B., Gonsalves, W., Maier, R., Silk, H., Stevens, N., Tysinger, J., & Wrightson, A. S. (2007). Smiles for Life: A National Oral Health Curriculum for Family Medicine. A model for curriculum development by STFM groups. Fam Med, 39(2), 88–90.

18. ElSalhy, M., Gill, M., Isaac, D. M., Littlechild, R., Baydala, L., & Amin, M. (2019). Integrating preventive dental care into general Paediatric practice for Indigenous communities: paediatric residents’ perceptions. INTERNATIONAL JOURNAL OF CIRCUMPOLAR HEALTH, 78(1), 1573162. 10.1080/22423982.2019.1573162

19. Gaffar, B., Bakhurji, E., AlKhateeb, R., AlHashim, H., AlGaoud, H., AlDaamah, Z., AlSaleh, J., Aldamanhori, R., AlHamid, S., AlBarrak, A., Siddiqui, I. A., & Virtanen, J. I. (2023). Exploring factors influencing nurses’ attitudes towards their role in dental care. PLOS ONE, 18(7), e0288927. 10.1371/journal.pone.0288927

20. Geiger, C. K., Kranz, A. M., Dick, A. W., Duffy, E., Sorbero, M., & Stein, B. D. (2019). Delivery of Preventive Oral Health Services by Rurality: A Cross Sectional Analysis. Journal of Rural Health, 35(1), 3–11. 10.1111/jrh.12340

21. Giles, E., Wray, F., Eskyte, I., Gray-Burrows, K. A., Owen, J., Bhatti, A., Zoltie, T., McEachan, R., Marshman, Z., Pavitt, S., West, R. M., & Day, P. F. (2022). HABIT: Health visitors delivering Advice in Britain on Infant Toothbrushing – an early-phase feasibility study of a complex oral health intervention. BMJ Open, 12(10), e059665. 10.1136/bmjopen-2021-059665

22. Gnaedinger, E. A. (2018). Fluoride varnish application, a quality improvement project implemented in a rural pediatric practice. Public Health Nursing, 35(6), 534–540. 10.1111/phn.12522

23. Goldstein, E. V., Dick, A. W., Ross, R., Stein, B. D., & Kranz, A. M. (2022). Impact of state level training requirements for medical providers on receipt of preventive oral health services for young children enrolled in Medicaid. Journal of Public Health Dentistry, 82(2), 156–165. 10.1111/jphd.12442

24. Gracner, T., Kranz, A. M., Li, K., Dick, A. W., & Geissler, K. (2023). The Patient Protection and Affordable Care Act and Pediatric Medical Clinicians’ Application of Fluoride Varnish. JAMA Network Open, 6(11), e2343087–e2343087. 10.1001/jamanetworkopen.2023.43087

25. Hachey, S., Clovis, J., & Lamarche, K. (2020). Strengthening the approach to oral health policy and practice in Canada. Paediatrics & Child Health, 25(2), 82–85. 10.1093/pch/pxz104

26. Harnagea, H., Couturier, Y., Shrivastava, R., Girard, F., Lamothe, L., Bedos, C. P., & Emami, E. (2017). Barriers and facilitators in the integration of oral health into primary care: a scoping review. BMJ Open, 7(9), Article e016078. 10.1136/bmjopen-2017-016078

27. Herndon, J. B., Tomar, S. L., Catalanotto, F. A., Vogel, W. B., & Shenkman, E. A. (2015). The effect of Medicaid primary care provider reimbursement on access to early childhood caries preventive services. Health Services Research, 50(1), 136–160. 10.1111/1475-6773.12200

28. Holve, S., Braun, P., Irvine, J. D., Nadeau, K., & Schroth, R. J. (2021). Early Childhood Caries in Indigenous Communities. Pediatrics, 147(6), 1–11. 10.1542/peds.2021-051481

29. Jackson, E. B. (2015). Outcomes of a Quality Improvement Project Examining Early Childhood Caries and Improving Identification of At Risk Patients in a Pediatric Medical Home Setting. JOURNAL OF PEDIATRIC NURSING-NURSING CARE OF CHILDREN & FAMILIES, 30(4), 543–549. 10.1016/j.pedn.2014.10.020

30. Johnson, S. C., & French, G. M. (2020). A Quality Improvement Project to Optimize Fluoride Varnish Use in a Pediatric Outpatient Clinic with Multiple Resident Providers. Hawaii J Health Soc Welf, *79*(5 Suppl 1), 7–12.

31. Kalhan, T. A., Un Lam, C., Karunakaran, B., Chay, P. L., Chng, C. K., Nair, R., Lee, Y. S., Fong, M. C. F., Chong, Y. S., Kwek, K., Saw, S. M., Shek, L., Yap, F., Tan, K. H., Godfrey, K. M., Huang, J., & Hsu, C. Y. S. (2020). Caries Risk Prediction Models in a Medical Health Care Setting. Journal of Dental Research, 99(7), 787–796. 10.1177/0022034520913476

32. Kim, P., Daly, J. M., Berkowitz, S., & Levy, B. T. (2020). Use of the Fluoride Varnish Billing Code in a Tertiary Care Center Setting. Journal of Primary Care & Community Health, 1–11. 10.1177/2150132720913736

33. Ko, A., Banks, J. T., Hill, C. M., & Chi, D. L. (2022). Fluoride Prescribing Behaviors for Medicaid-Enrolled Children in Oregon. American Journal of Preventive Medicine, 62(2), e69–e76. 10.1016/j.amepre.2021.06.016

34. Kranz, A. M., Lee, J., Divaris, K., Baker, A. D., & Vann, W., Jr. (2014a). North Carolina physician-based preventive oral health services improve access and use among young Medicaid enrollees. Health Aff (Millwood), 33(12), 2144–2152. 10.1377/hlthaff.2014.0927

35. Kranz, A. M., Opper, I. M., Stein, B. D., Ruder, T., Gahlon, G., Sorbero, M., & Dick, A. W. (2022). Medicaid Payment and Fluoride Varnish Application During Pediatric Medical Visits. Med Care Res Rev, 79(6), 834–843. 10.1177/10775587221074766

36. Kranz, A. M., Preisser, J. S., & Rozier, R. G. (2015). Effects of Physician-Based Preventive Oral Health Services on Dental Caries. Pediatrics, 136(1), 107–114. 10.1542/peds.2014-2775

37. Kranz, A. M., Rozier, G., Preisser, J. S., Stearns, S. C., Weinberger, M., & Lee, J. Y. (2014b). Comparing Medical and Dental Providers of Oral Health Services on Early Dental Caries Experience. American Journal of Public Health, 104(7), e92–99. 10.2105/AJPH.2014.301972

38. Kranz, A. M., Rozier, R. G., Stein, B. D., & Dick, A. W. (2020). Do Oral Health Services in Medical Offices Replace Pediatric Dental Visits? Journal of Dental Research, 99(7), 891–897. 10.1177/0022034520916161

39. Kressin, N. R., Nunn, M. E., Singh, H., Orner, M. B., Pbert, L., Hayes, C., Culler, C., Glicken, S. R., Palfrey, S., Geltman, P. L., Cadoret, C., Henshaw, M. M., Kressin, N. R., Nunn, M. E., Singh, H., Orner, M. B., Pbert, L., Hayes, C., Culler, C., & Glicken, S. R. (2009). Pediatric clinicians can help reduce rates of early childhood caries: effects of a practice based intervention. Medical Care, 47(11), 1121–1128. 10.1097/MLR.0b013e3181b58867

40. Krol, D. M., & Whelan, K. (2023). Maintaining and Improving the Oral Health of Young Children. Pediatrics, 151(1), 1–8. 10.1542/peds.2022-060417

41. Kyoon-Achan, G., Schroth, R. J., DeMare, D., Sturym, M., Edwards, J. M., Sanguins, J., Campbell, R., Chartrand, F., Bertone, M., & Moffatt, M. E. K. (2021). First Nations and Metis peoples’ access and equity challenges with early childhood oral health: a qualitative study. Int J Equity Health, 20(1), 134. 10.1186/s12939-021-01476-5

42. Levac, D., Colquhoun, H., & O’Brien, K. K. (2010). Scoping studies: advancing the methodology. Implementation Science, 5(1), 69. 10.1186/1748-5908-5-69

43. Lienhart, G., Elsa, M., Farge, P., Schott, A. M., Thivichon-Prince, B., & Chaneliere, M. (2023). Factors perceived by health professionals to be barriers or facilitators to caries prevention in children: a systematic review. BMC Oral Health, 23(1), 767. 10.1186/s12903-023-03458-1

44. Long, C. M., Quinonez, R. B., Beil, H. A., Close, K., Myers, L. P., Vann, W. F., & Rozier, R. G. (2012). Pediatricians’ assessments of caries risk and need for a dental evaluation in preschool aged children. BMC Pediatrics, 12(1), 49–49. 10.1186/1471-2431-12-49

45. Lukac, P. J., Bell, D., Sreedharan, P., Gornbein, J. A., & Lerner, C. (2023). The Application of Dental Fluoride Varnish in Children: A Low Cost, High-Value Implementation Aided by Passive Clinical Decision Support. Appl Clin Inform, 14(2), 245–253. 10.1055/a-2011-8167

46. Maher, L., Phelan, C., Lawrence, G., Torvaldsen, S., Dawson, A., & Wright, C. (2012). The Early Childhood Oral Health Program: promoting prevention and timely intervention of early childhood caries in NSW through shared care. Health Promotion Journal of Australia, 23(3), 171–176.

47. McCulley, M. G., Prihoda, K., & Ayres, C. (2022). Improving the Quality of Oral Health Screening for Young Children in Primary Care. Journal of Nursing Practice Applications & Reviews of Research, 12(2), 41–50. 10.13178/jnparr.2022.12.02.1206

48. Melgar, X. C., Azanedo, D., & Hugo, F. N. (2024). Towards the integration of prevention and control of oral diseases within child primary healthcare: The case of Peru. Community Dent Oral Epidemiol, 52(4), 509–517. 10.1111/cdoe.12945

49. Meyer, B. D., & Danesh, D. O. (2021). The Impact of COVID-19 on Preventive Oral Health Care During Wave One [Perspective]. FRONTIERS IN DENTAL MEDICINE, Volume 2 – 2021. 10.3389/fdmed.2021.636766

50. Moher, D., Liberati, A., Tetzlaff, J., & Altman, D. G. (2009). Preferred reporting items for systematic reviews and meta-analyses: the PRISMA statement. PLoS Med, 6(7), e1000097. 10.1371/journal.pmed.1000097

51. Murphy, K. L., & Larsson, L. S. (2017). Interprofessional oral health initiative in a nondental, American Indian setting. Journal of the American Association of Nurse Practitioners, 29(12), 733–740. 10.1002/2327-6924.12517

52. Neumann, A. S., Lee, K. J., Gussy, M. G., Waters, E. B., Carlin, J. B., Riggs, E., & Kilpatrick, N. M. (2011). Impact of an oral health intervention on pre-school children <3 years of age in a rural setting in Australia. JOURNAL OF PAEDIATRICS AND CHILD HEALTH, 47(6), 367–372. 10.1111/j.1440-1754.2010.01988.x

53. Okah, A., Williams, K., Talib, N., & Mann, K. (2018). Promoting Oral Health in Childhood: A Quality Improvement Project. Pediatrics, 141(6). 10.1542/peds.2017-2396

54. Okunseri, C., Szabo, A., Garcia, R. I., Jackson, S., & Pajewski, N. M. (2010). Provision of fluoride varnish treatment by medical and dental care providers: variation by race/ethnicity and levels of urban influence. Journal of Public Health Dentistry, 70(3), 211–219. 10.1111/j.1752-7325.2010.00168.x

55. Okunseri, C., Szabo, A., Jackson, S., Pajewski, N. M., Garcia, R. I., Okunseri, C., Szabo, A., Jackson, S., Pajewski, N. M., & Garcia, R. I. (2009). Increased children’s access to fluoride varnish treatment by involving medical care providers: effect of a Medicaid policy change. Health Services Research, 44(4), 1144–1156. 10.1111/j.1475-6773.2009.00975.x

56. Olatosi, O. O., Schroth, R. J., DeMaré, D., Manigque, M., Mittermuller, B., Edwards, J., Yerex, K., Wong, P. D., Lavoie, J., Sanguins, J., Chelikani, P., Nicolae, A., Lamoureux, J., Campbell, R., Bertone, M., & Amin, M. (2025a). Identifying training needs of healthcare providers to implement caries risk assessment [Original Research]. FRONTIERS IN ORAL HEALTH, Volume 6 – 2025. 10.3389/froh.2025.1641307

57. Olatosi, O. O., Schroth, R. J., DeMaré, D., Mittermuller, B., Manigque, M., Edwards, J., Amin, M. S., Nicolae, A., Lavoie, J., Sanguins, J., Chelikani, P., Wong, P. D., Lamoureux, J., Bertone, M., Yerex, K., Campbell, R., & The Working Together for early childhood oral health study, t. (2025b). Healthcare providers’ perspectives on the Canadian Caries Risk Assessment Tool implementation in Indigenous pediatric primary care: a qualitative study. BMC Oral Health, 25(1), 708. 10.1186/s12903-025-06036-9

58. Olatosi, O. O., Schroth, R. J., DeMare, D., Mittermuller, B. A., Manigque, M., Edwards, J., Amin, M., Nicolae, A., Lavoie, J., Sanguins, J., Chelikani, P., Wong, P., Lamoureux, J., Bertone, M., Yerex, K., Campbell, R., & Working Together for Early Childhood Oral Health Study, T. (2025c). Recommendations for Integrating Caries Risk Assessment into Primary Care for Indigenous Children. JDR Clin Trans Res, 23800844251372545. 10.1177/23800844251372545

59. Pahel, B. T., Rozier, R. G., & Stearns, S. C. (2010). Agreement between structured checklists and Medicaid claims for preventive dental visits in primary care medical offices. Health Informatics Journal, 16(2), 115–128. 10.1177/1460458210364036

60. Pahel, B. T., Rozier, R. G., Stearns, S. C., & Quinonez, R. B. (2011). Effectiveness of preventive dental treatments by physicians for young Medicaid enrollees. Pediatrics, 127(3), e682– 689. 10.1542/peds.2010-1457

61. Patton, S., & Severe, S. (2020). Nursing Students’ Assessment and Parent Reports of Their Children’s Oral Health Behaviors as Predictors of Tooth Decay Risk—A Cross-Sectional, Correlational Study. JOURNAL OF ADVANCED ORAL RESEARCH, 11(1), 45–51. 10.1177/2320206819895846

62. Quinonez, R. B., Kranz, A. M., Lewis, C. W., Barone, L., Boulter, S., O’Connor, K. G., & Keels, M. A. (2014). Oral Health Opinions and Practices of Pediatricians: Updated Results From a National Survey. Academic Pediatrics, 14(6), 616–623. 10.1016/j.acap.2014.07.001

63. Rabiei, S., Mohebbi, S. Z., Yazdani, R., & Virtanen, J. I. (2014). Primary care nurses’ awareness of and willingness to perform children’s oral health care. BMC Oral Health, 14, 26. 10.1186/1472-6831-14-26

64. Rolnick, S. J., Jackson, J. M., DeFor, T. A., & Flottemesch, T. J. (2015). Fluoride Varnish Application in the Primary Care Setting. A Clinical Study. Journal of Clinical Pediatric Dentistry, 39(4), 311–314. 10.17796/1053-4628-39.4.311

65. Roth, L. T., Robbins-Milne, L., Sirota, D., & Lane, M. (2020). A Resident-Led QI Project to Improve Dental Health at a Primary Care Pediatric Practice. J Grad Med Educ, 12(5), 571–577. 10.4300/jgme-d-19-00959.1

66. Rowan-Legg, A., & Canadian Paediat, S. (2013). Oral health care for children – a call for action. Paediatrics & Child Health, 18(1), 37–43. 10.1093/pch/18.1.37

67. Rozier, R. G., Stearns, S. C., Pahel, B. T., Quinonez, R. B., & Park, J. (2010). How A North Carolina Program Boosted Preventive Oral Health Services For Low-Income Children. Health Affairs, 29(12), 2278–2285. 10.1377/hlthaff.2009.0768

68. Schroth, R. J., Quinonez, C., Shwart, L., & Wagar, B. (2016). Treating Early Childhood Caries under General Anesthesia: A National Review of Canadian Data. J Can Dent Assoc, 82, g20.

69. Schroth, R. J., Rothney, J., Sturym, M., Dabiri, D., Dabiri, D., Dong, C. C., Grant, C. G., Kennedy, T., & Sihra, R. (2021). A systematic review to inform the development of a Canadian caries risk assessment tool for use by primary healthcare providers. Int J Paediatr Dent, 31(6), 767–791. 10.1111/ipd.12776

70. Sibley, J. A. (2018). Cost-Benefit Analysis of Providing Fluoride Varnish in a Pediatric Primary Care Office. J Pediatr Health Care, 32(6), 620–626. 10.1016/j.pedhc.2018.05.007

71. Silk, H., Sachs Leicher, E., Alvarado, V., Cote, E., & Cote, S. (2018). A multi-state initiative to implement pediatric oral health in primary care practice and clinical education. Journal of Public Health Dentistry, 78(1), 25–31. 10.1111/jphd.12225

72. Sudhanthar, S., Lapinski, J., Turner, J., Gold, J., Sigal, Y., Thakur, K., Napolova, O., & Stiffler, M. (2019). Improving oral health through dental fluoride varnish application in a primary care paediatric practice. BMJ Open Qual, 8(2), e000589. 10.1136/bmjoq-2018-000589

73. Taylor, E., Marino, D., Thacker, S., DiMarco, M., Huff, M., & Biordi, D. (2014). Expanding oral health preventative services for young children: a successful interprofessional model. J Allied Health, 43(1), e5–9.

74. Tungare, S., & Paranjpe, A. G. (2025). Early Childhood Caries. In StatPearls.

75. Turner, S., Brewster, L., Kidd, J., Gnich, W., Ball, G. E., Milburn, K., Pitts, N. B., Goold, S., Conway, D. I., & Macpherson, L. M. (2010). Childsmile: the national child oral health improvement programme in Scotland. Part 2: Monitoring and delivery. Br Dent J, 209(2), 79–83. 10.1038/sj.bdj.2010.629

76. Turton, B., Durward, C., Crombie, F., Sokal Gutierrez, K., Soeurn, S., & Manton, D. J. (2021). Evaluation of a community based early childhood caries (ECC) intervention in Cambodia. Community Dentistry & Oral Epidemiology, 49(3), 275–283. 10.1111/cdoe.12599

77. Verlinden, D. A., Schuller, A. A., Vermaire, J. H., & Reijneveld, S. A. (2024). Referral from well child care clinics to dental clinics leads to earlier initiation of preventive dental visits: A quasi experimental study. International Journal of Paediatric Dentistry, 34(2), 190–197. 10.1111/ipd.13124

78. Veschusio, C. N., Probst, J. C., Martin, A. B., Hardin, J. W., & L. Hale, N. (2016). Impact of South Carolina’s Medicaid fluoride varnish reimbursement policy on children’s receipt of fluoride varnish in medical and dental settings. Journal of Public Health Dentistry, 76(4), 356–361. 10.1111/jphd.12163

79. Wigen, T. I., & Wang, N. J. (2017). Referral of young children to dental personnel by primary care nurses. International Journal of Dental Hygiene, 15(3), 249–255. 10.1111/idh.12238

80. Zea, A., & Henshaw, M. (2022). Promoting Children’s Health Equity With Medical-Dental Integration. AMA J Ethics, 24(1), E33–40. 10.1001/amajethics.2022.33

